# Identifying and Forecasting Importation and Asymptomatic Spreaders of Multi-drug Resistant Organisms in Hospital Settings

**DOI:** 10.1101/2024.07.14.24310393

**Authors:** Jiaming Cui, Jack Heavey, Eili Klein, Gregory R. Madden, Costi D. Sifri, Anil Vullikanti, B. Aditya Prakash

## Abstract

Healthcare-associated infections (HAIs) from multi-drug resistant organisms (MDROs) pose a significant challenge for healthcare systems. Patients can arrive at hospitals already infected (“importation”) or acquire infections during their stay (“nosocomial infection”). Many cases, often asymptomatic, complicate rapid identification due to testing limitations and delays. Although recent advancements in mathematical modeling and machine learning have aimed to identify at-risk patients, these methods face challenges: transmission models often overlook valuable electronic health record (EHR) data, while machine learning approaches typically lack mechanistic insights into underlying processes. To address these issues, we propose NeurABM, a novel framework that integrates neural networks and agent-based models (ABM) to leverage the strengths of both methods. NeurABM simultaneously learns a neural network for patient-level importation predictions and an ABM for infection identification. Our findings show that NeurABM significantly outperforms existing methods, marking a breakthrough in accurately identifying importation cases and forecasting future nosocomial infections in clinical practice.

## Introduction

Healthcare-associated infections (HAIs), especially those caused by multi-drug resistant organisms (MDROs), pose a significant threat to patient safety and burden the healthcare system with increased costs due to longer hospital stays and more expensive therapies [46, 42, 40, 48, 18, 27]. Approximately 3% of hospitalized patients in the United States acquire an HAI during their stay, resulting in more than 35,000 deaths annually [7, 23, 14, 38]. In many occasions, patients may have already been colonized (pathogens present on patients without causing disease [31]) or infected but may be asymptomatic at admission (i.e., importation cases) [13]. For instance, the European Centre for Disease Prevention and Control (ECDC) estimates that importation cases contribute to 13% of HAI cases in Germany and 18.9% in Spain [47]. These importation cases can spread HAI-causing pathogens either directly via patient-patient contacts or indirectly via healthcare workers (HCWs) and contaminated physical surfaces [5, 4, 45] and lead to nosocomial infection cases, which can also be asymptomatic but further spread pathogens to additional healthy patients [44].

Despite the critical concerns associated with importation and nosocomial infection cases, identifying these cases rapidly and accurately still remains a challenging problem. Current methods to identify them include surveillance tests [35, 28], machine learning-based methods [37, 22], and transmission modeling-based methods [34, 20, 3, 32, 30, 49, 16]. However, each of these methods suffers from unavoidable drawbacks. For example, surveillance tests, such as culture or PCR tests, are common in hospitals; however, they are costly, require time to process, and are not 100% accurate [12]. Additionally, they are not applicable for all MDROs; typically, they can be used for only a subset of MDROs [1]. Machine learning and statistical techniques use patients’ electronic health record (EHR) data to predict the probability of importation and nosocomial infection cases [37, 22] (see Figure 1a). However, the performance of machine learning methods has not proven to be sufficiently robust for clinical practice due to many reasons, including imbalance (since HAI cases form a very small fraction of the entire patient population) and bias in the data (since testing is generally not done in a systematic manner). Moreover, explicitly incorporating physical epidemiological mechanisms into machine learning frameworks remains an open problem. ML frameworks tend to focus on data-driven correlations rather than understanding and using complex causal relationships and dynamics of disease transmission [39]. Finally, modeling-based methods are based on detailed mechanistic models (e.g., compartmental mixing models [30, 49] and agent-based models (ABMs) [3, 34, 20]) to capture the transmission dynamics of HAIs within a healthcare facility. They are calibrated to infections in the hospital and use projections from such models for prediction. Although ABMs have used information about contact networks between patients and providers within healthcare facilities to model the infection status of an individual patient, they still cannot directly incorporate the risk factor of each patient from the EHR data, such as medications, lab results, vital signs, and device use history into modelling. Besides, they rely on contact networks to run simulations, which makes their forecasts for future nosocomial infections less reliable since people have to make assumptions on future contact networks to run the model (see Figure 1b). As we will also show in the later results section, these limitations lead to suboptimal performance in identifying importation and forecasting future nosocomial infection cases.

**Figure 1:**
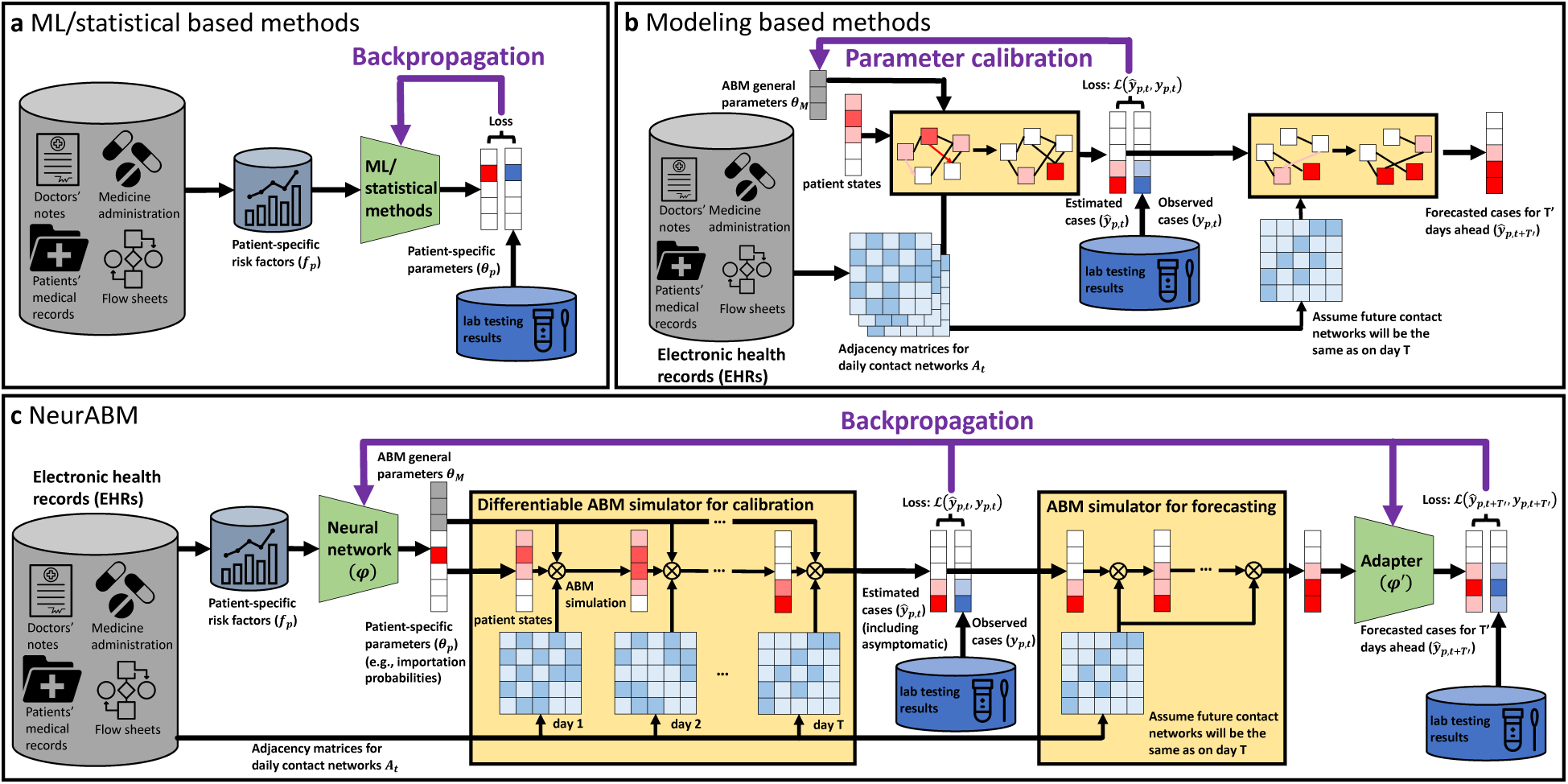
NeurABM framework. **a)** Traditional machine learning and statistical methods use patients’ risk factors to predict the probability of being importation and/or future nosocomial infection cases. It is hard to explicitly incorporate epidemiological mechanisms. **b)** Modeling based methods are built on mechanistic models to capture the HAI spread in healthcare facilities. Here, observed cases are used to train an ABM to (1) learn underlying ABM parameters and (2) pin the ABM states to the spread dynamics until today, which allows people to run ABM simulations for *T′* more days for forecasting. However, they cannot leverage the the risk factor of each patient from the EHR and have to make assumptions for future contact networks. **c)** To address this issue, we propose NeurABM framework to couple both neural networks and ABM simultaneously. Specifically, it is composed of two main components: the neural network component and the agent-based model (ABM) simulator component. The neural network component takes patient-specific risk factors data collected from EHR as input and outputs both the ABM parameters (applicable to every patient) and patient-specific parameters (importation probabilities in this paper). The ABM simulator component takes daily contact networks collected from EHR and the parameters as input, runs simulations for *T* days, and outputs the probability that each patient is in the *Carriage* state for each day. We then compare them with the ground-truth observations (via lab testing) and compute the first loss. To forecast future nosocomial infections, we start from predictions on day *T* and run simulations for *T′* more days. Note that these *T′* more days are for future, we assume that the contact networks will be the same as on day *T*, and use another adapter network to correct the potential bias caused by this assumption and get the final forecast. We also compute the loss of this forecast with ground-truth observations. With both losses, we backpropagate the total loss to adjust neural network parameters. This design allows us to train both the neural network and the ABM simultaneously in an end-to-end manner, mitigating the issues encountered when using either component individually.

From a broader perspective, improved identification of importation cases and forecasts of future noso-comial infections could also improve hospital management. Practices to prevent the spread of MDROs within hospitals, such as testing, quarantine, and isolation, consume limited healthcare resources and could only be implemented on a small group of patients. Meanwhile, decisions about prioritizing limited re-sources are critical to ensure effective and efficient delivery of healthcare services [41], where agent-based models [11, 19, 24, 26] and machine learning [25] have proven to be invaluable tools for optimizing these decisions.

In this work, we propose a new framework, NeurABM, to identify HAI importation and forecast future nosocomial infection cases by *coupling* a neural network and an ABM and training simultaneously. We use methicillin-resistant *Staphylococcus aureus* (MRSA) as an example HAI in later sections. Figure 1 shows an overview of our framework. As shown in Figure 1c, the neural network *ϕ* estimates the importation probability for each patient using EHR data, while the ABM incorporates MRSA dynamics and is used to estimate the MRSA infection probability. After training, NeurABM runs as a discrete time process; at each time step *t*, the following two steps are performed: (1) the neural network estimates the importation probability (i.e., identifies importation cases) for each new patient who enters the hospital, (2) the ABM (which keeps track of the disease states of all patients in hospital till time *t −* 1) runs the next step of the disease simulation to estimate all disease states (i.e., identify nosocomial infection cases, including those which are asymptomatic) at time *t*. When forecasting for future, we assume that the contact networks will be the same as on day *T*, and use another adapter network *ϕ′* to learn and mitigate the bias caused by this assumption. The parameters of NeurABM consist of two parts: those of the neural network and those of the ABM (e.g., general parameters such as transmission and recovery rates, and patient-specific parameters such as importation probabilities in this paper); these parameters are learned by minimizing a loss function that considers the errors in the ABM projections and ground truth incidence data from EHR. Since the dynamics of MRSA transmission depend on the importation model (and conversely, through this kind of training process), this approach couples the neural network and ABM and is trained end-to-end, which mitigates the issues in using either of them individually. NeurABM significantly extends the work of Chopra and Rodriguez et al. [9], which was the first method to consider such a joint deep learning and ABM approach, by introducing approximation techniques to scale the disease model, incorporating rich patient-level EHR data, and new techniques to train the pipeline robustly.

We demonstrate the performance of NeurABM using EHR data for patients at the University of Virginia (UVA) hospital intensive care units (ICUs). Our results show that NeurABM not only identifies importation cases but also forecasts future nosocomial infection cases better than other machine learning or modeling-based baselines. Note that the NeurABM is a general framework that integrates both neural networks and mechanistic models in an end-to-end way, one which can be easily extended to other ABMs or EHR data and study other clinical problems.

## Results

EHR data was used from the UVA hospital to construct patient contact networks (used by the ABM) and collect patient risk factors (used by the neural network). We use the SIS-ABM model [21, 10] as the ABM for disease transmission in NeurABM. Ground-truth MRSA infections are identified from lab test results for each patient in the EHR. For each week *k*, contact networks, patient risk factors, and lab test results until week *k −* 1 were used to train the NeurABM and identify importation cases before week *k −* 1. We then ran the SIS-ABM model for 7 more days to infer the infection states of patients for week *k*. Two setups were assessed: (1) When identifying current nosocomial infection cases in hospital, contact networks in week *k* are used in this process, similar to the approach discussed in previous work [34] for detection of asymptomatic cases, although their ABM is different. They also did not consider the importation problem. (2) When forecasting future nosocomial infection cases, no data in week *k* is used, which exactly follows a real-world forecasting setup that no future information is known. For evaluation, we compare NeurABM with a broad range of baselines in machine learning categories (feedforward neural network [33], decision tree [17], naive bayes [6], XGBoost [8], and Autoencoder+KNN [36, 15]), modeling categories (the SIS-ABM model [21, 10], SILI-ABM model [34]), and clinical heuristic categories (length of stay [34]). Note that since the SILI-ABM model [34] is not designed to identify importation cases, we only compare it for identifying nosocomial infection cases.

### Identifying importation cases

In Figure 2a, we show the precision-recall curves [17] for NeurABM and other methods. Intuitively, precision measures the accuracy of positive predictions, while recall measures the ability to identify all actual positives. Note that in clinical practice, very low precision is not very useful, since this means too many tests and treatments do not help to identify and treat MRSA cases. Therefore, we always expect high recall with not-too-low precision. Following previous work [29], we consider precision smaller than 0.25 as clinically inapplicable and focus on three important precision levels: 0.25, 0.5, and 0.75 (dashed grey lines). NeurABM consistently achieves the highest recall at precision levels of 0.25, 0.5, and 0.75, demonstrating the effectiveness of the NeurABM framework. For example, a precision of 0.25 means that, on average, one out of four patients identified as importation cases are actually importation cases. Additionally, when precision is 0.25, NeurABM achieves 0.74 recall, while the best baseline (feedforward neural network) only achieves a recall of 0.40. This means that NeurABM could identify 55 out of 74 importation cases in UVA ICU, while the feedforward neural network could only identify 29 out of 74, missing 26 more patients than NeurABM. Finally, the area under the precision-recall curve (AUPRC) for NeurABM is the largest (0.60) among all methods. In Figure 2b, we show how the negative predictive value (NPV) changes with threshold changes (when the estimated probability is higher than the threshold, we classify this patient as a MRSA importation case, and vice versa). The NPV is the fraction of the number of true negative cases over predicted negative cases. Intuitively, a higher NPV means that when we predict a patient as negative, he or she is less likely to be a false negative patient that we fail to identify. We can see that NeurABM’s NPV rate is always higher than 0.9 and other baselines, indicating that NeurABM can identify the importation cases well with fewer missing/undetected patients. Figure 2c presents the receiver operating characteristic (ROC) curve [17], which illustrates the trade-off between the true positive rate (the proportion of actual positives correctly identified, which is recall) and the false positive rate (the proportion of negatives incorrectly classified as positives, which is 1-specificity) across different classification thresholds. Here, the area under the curve (AUC-ROC) for our framework is still the largest (0.86) compared to other baselines. Intuitively, an AUC-ROC of 0.86 means that, on average, there is an 86% probability that NeurABM will predict higher importation probabilities for an importation case compared to a non-importation case. In the table in Figure 2d, we also list the recall, F1 score, and false positive rate corresponding to different precision values. NeurABM consistently achieves the highest recall and F1 score with a given precision, indicating the effectiveness of our framework in identifying MRSA importation cases.

**Figure 2:**
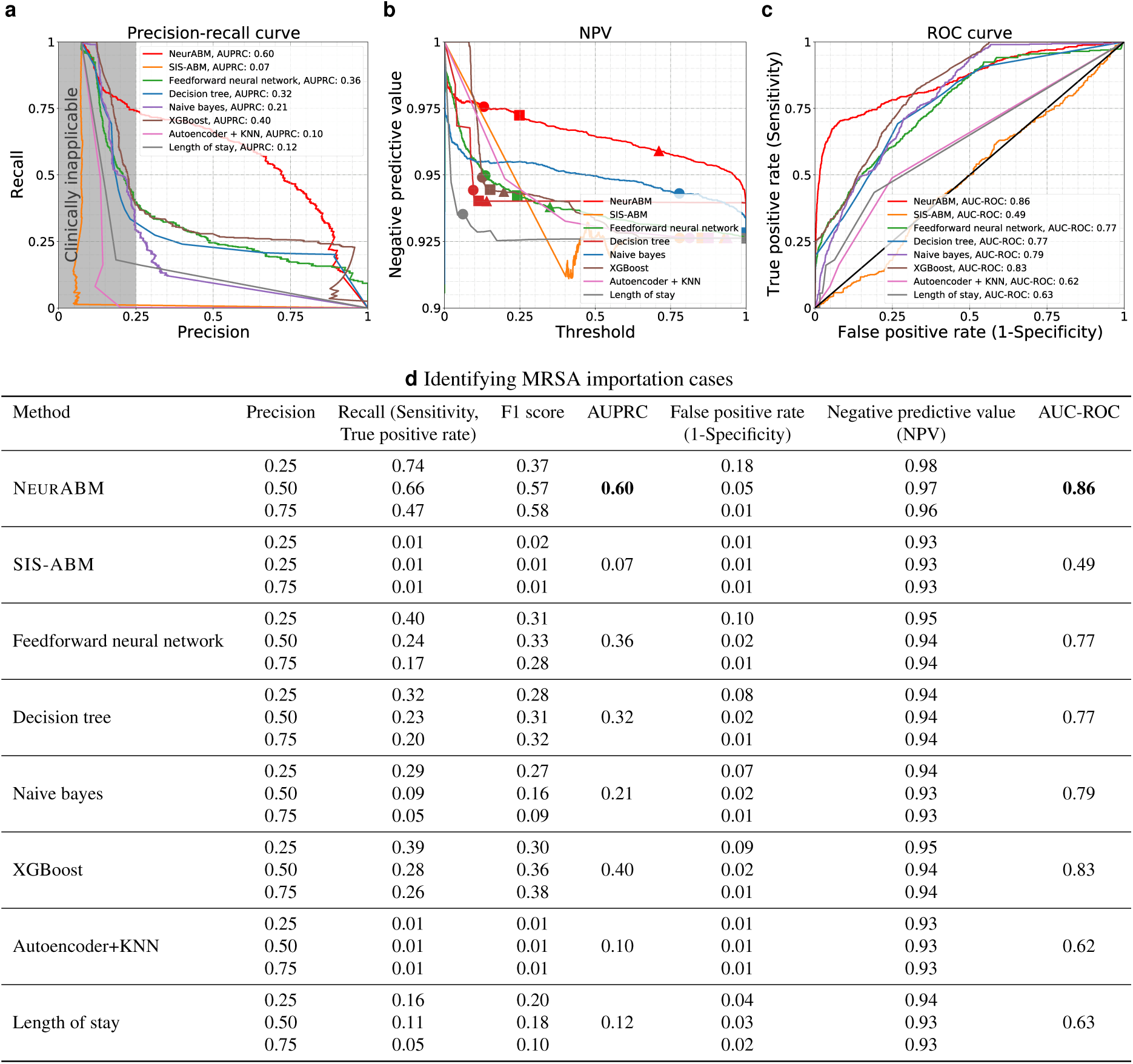
The performance in identifying importation cases. **a)** The precision-recall curves (PRC). The x-axis represents precision, and the y-axis represents recall. The red and other color curves represent NeurABM and other baselines. A larger area under the precision-recall curve (AUPRC) indicates better performance. AUPRC values are listed in the legends, and NeurABM has the highest AUPRC value. **b)** The negative predictive value (NPV) with different thresholds. The x-axis is the threshold for classification, and the y-axis is the NPV value. Circles, squares, and triangles correspond to the thresholds and NPV values where precision is 0.25, 0.5, and 0.75, respectively. A higher NPV value indicates fewer missing importation cases that are not identified and therefore better performance, and NeurABM has the highest NPV values. **c)** The receiver operating characteristic (ROC) curves in identifying MRSA importation cases. The x-axis is the false positive rate, and the y-axis is the true positive rate. A larger area under the ROC (AUC-ROC) indicates better performance. AUC-ROC values are listed in the legends, and NeurABM has the highest AUC-ROC value. **d)** The recall, F1 score, AUPRC, false positive rate, NPV, and AUC-ROC under different precisions (0.25, 0.5, 0.75). The best AUPRC and AUC-ROC are in bold.

### Identifying current nosocomial infection cases

Next, we assessed the effectiveness of NeurABM to identify current nosocomial infection cases of MRSA. As shown in Figure 3a, the x-axis and y-axis represent precision and recall, respectively. The red curve represents the results for NeurABM. As shown in the figure, the area under the precision-recall curve for our framework is the largest (0.69) compared to other baselines. The dashed grey lines correspond to the precision of 0.25, 0.5, and 0.75. Again, NeurABM consistently achieves the highest recall with precision equal to 0.25, 0.5, and 0.75, indicating that our framework is effective. For example, a precision of 0.5 means that, on average, one out of two patient days we identified are actual nosocomial infection patient days. Additionally, when precision is 0.5, NeurABM achieves 0.81 recall, while the best baseline (XGBoost) only achieves a recall of 0.60. This means that NeurABM was able to identify 331 out of 408 nosocomial infection patient days in UVA ICU, while XGBoost could only identify 245 out of 408, missing 86 more patient days than NeurABM. In Figure 3b, we show how the NPV changes with the threshold for classification. We can see that the NPV rate is always higher than 0.9 and other baselines, indicating that NeurABM can identify current nosocomial infection cases well with fewer missing/undetected patients. The ROC curve in Figure 3c demonstrates that the AUC-ROC for the NeurABM framework is the highest (0.88) compared to other baselines. Intuitively, this means there is an 88% probability that NeurABM will assign higher probabilities to actual current nosocomial infection cases compared to non-nosocomial infection cases. In the table in Figure 3d, NeurABM always achieves the highest recall and F1 score with a given precision, demonstrating the effectiveness of our framework in identifying current nosocomial MRSA infection cases.

**Figure 3:**
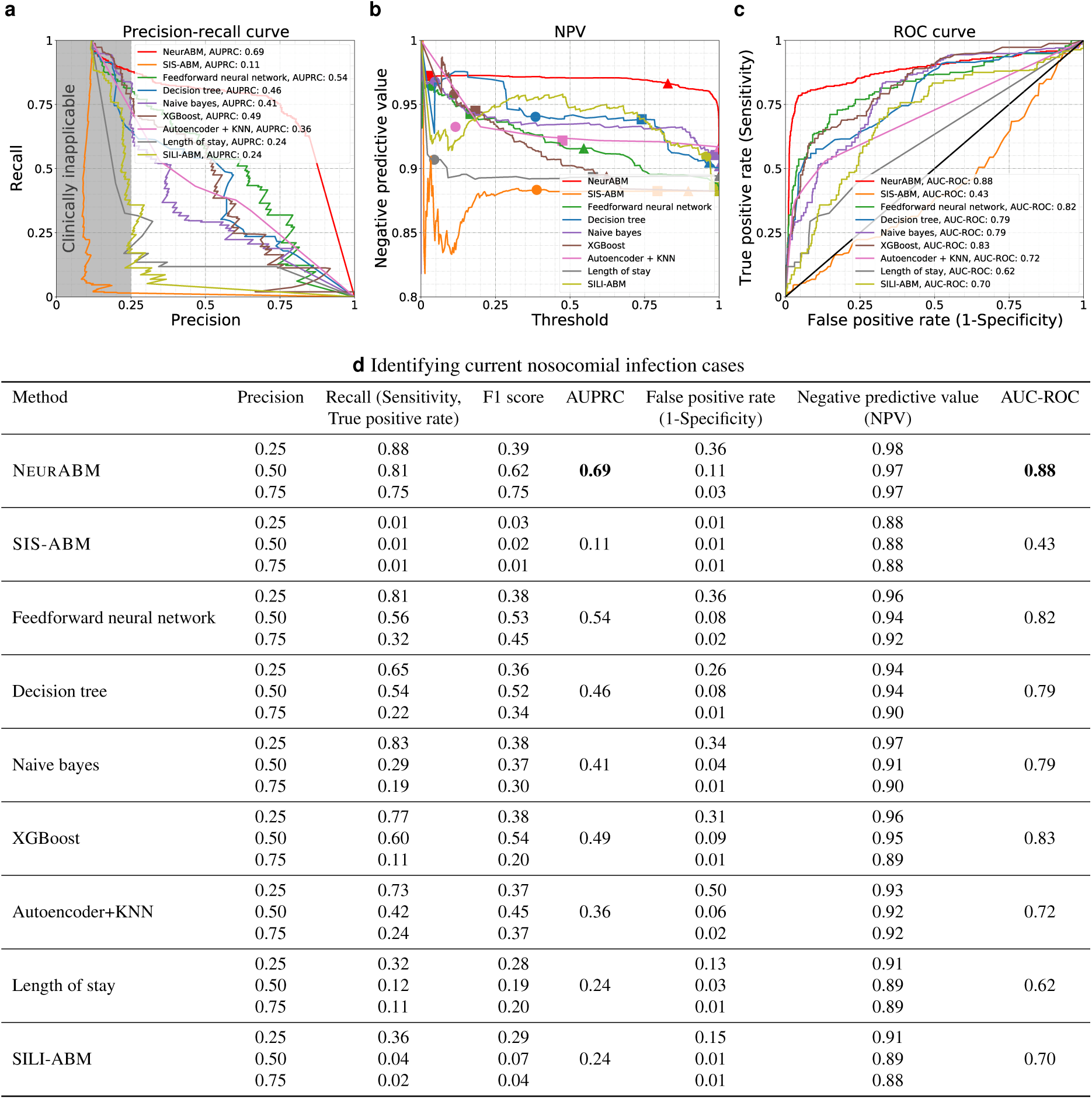
The performance in identifying current nosocomial infection cases. **a)** The precision-recall curves. The red and other color curves represent NeurABM and other baselines. Higher AUPRC is better, and NeurABM has the highest AUPRC value. **b)** The negative predictive value with different thresholds. Circles, squares, and triangles correspond to the thresholds and NPV values where precision is 0.25, 0.5, and 0.75, respectively. Higher NPV value is better, and NeurABM has the highest NPV values. **c)** The receiver operating characteristic curves in identifying MRSA nosocomial infection cases. Higher AUC-ROC is better, and NeurABM has the highest AUC-ROC value. **d)** The recall, F1 score, AUPRC, false positive rate, NPV, and AUC-ROC under different precisions. The best AUPRC and AUC-ROC are in bold.

### Forecasting future nosocomial infection cases

We then directed our attention to examining the ability of NeurABM to identify current nosocomial infection cases of MRSA. As shown in Figure 3a, the x-axis and y-axis represent precision and recall, respectively. The red curve represents the results for NeurABM. As shown in the figure, the area under the precision-recall curve for our framework is the largest (0.85) compared to other baselines. The dashed grey lines correspond to the precision of 0.25, 0.5, and 0.75. Again, NeurABM always achieves the highest recall with precision equal to 0.25, 0.5, and 0.75, indicating that our framework is effective. For example, when precision is 0.75, NeurABM achieves 0.88 recall, while the best baseline (Autoencoder+KNN) only achieves a recall of 0.5. This means that we could forecast 275 out of 312 future nosocomial infection patient days in UVA ICU, while the Autoencoder+KNN could only forecast 156 out of 312, missing 119 more patient days than NeurABM. In Figure 4b, we show how the NPV changes with the threshold for classification. We can see that the NPV rate is always higher than 0.9 and other baselines, indicating that NeurABM can forecast future nosocomial infection cases well with fewer missing/undetected patients. In Figure 4c, the AUC-ROC for NeurABM framework is the largest (0.92) compared to other baselines. Intuitively, this means there is an 92% probability that NeurABM will assign higher probabilities to future nosocomial infection cases compared to non-nosocomial infection cases. In the table in Figure 4d, NeurABM always achieves the highest recall and F1 score with a given precision, demonstrating the effectiveness of our framework in fore-casting future nosocomial MRSA infection cases. We also ran experiments to forecast for 2 weeks ahead (see Supplementary Information), and the results show that NeurABM still performs better than other baselines.

**Figure 4:**
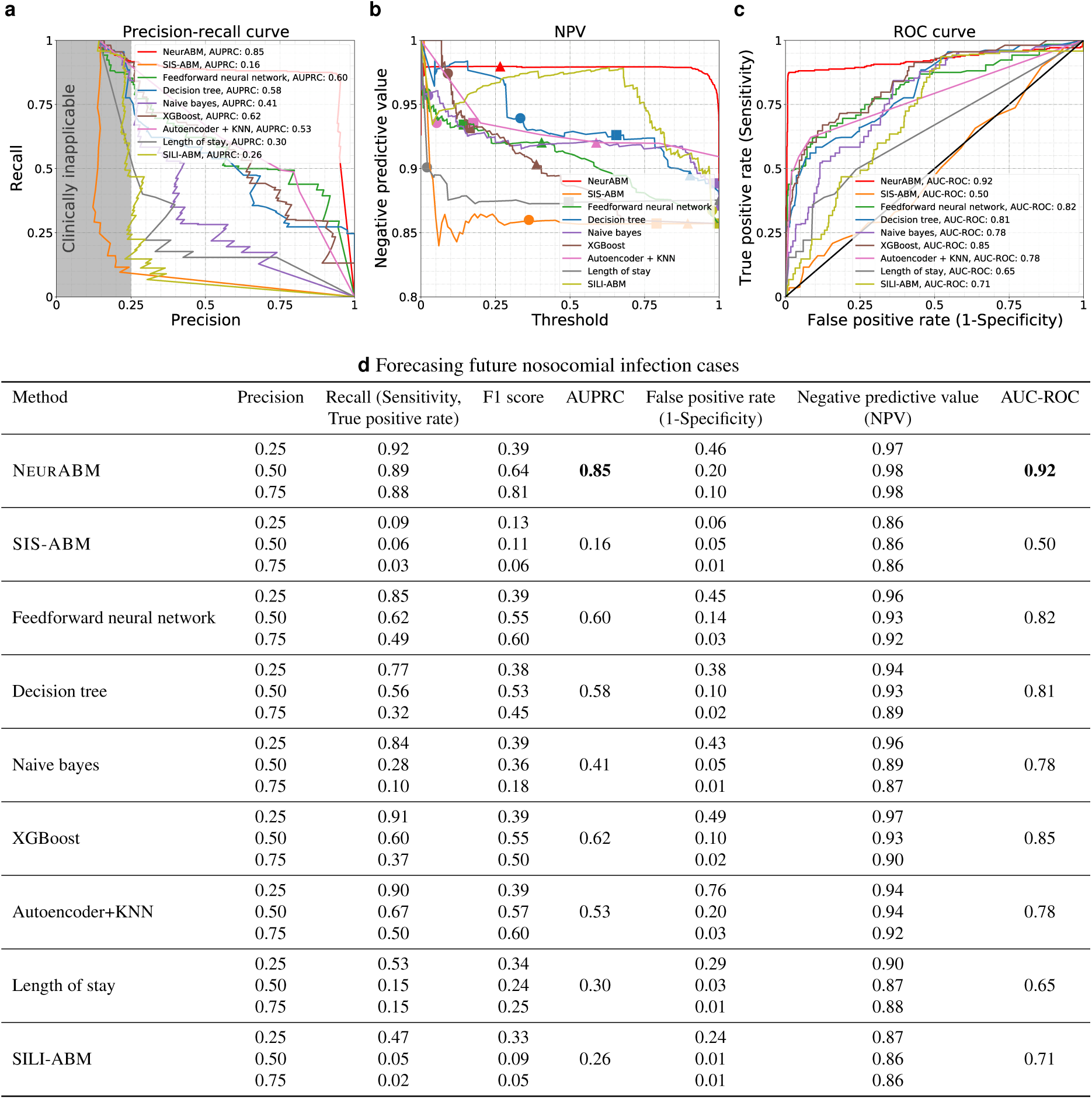
The performance in forecasting future nosocomial infection cases. **a)** The precision-recall curves. The red and other color curves represent NeurABM and other baselines. Higher AUPRC is better, and NeurABM has the highest AUPRC value. **b)** The negative predictive value with different thresholds. Circles, squares, and triangles correspond to the thresholds and NPV values where precision is 0.25, 0.5, and 0.75, respectively. Higher NPV value is better, and NeurABM has the highest NPV values. **c)** The receiver operating characteristic curves in forecasting future MRSA nosocomial infection cases. Higher AUC-ROC is better, and NeurABM has the highest AUC-ROC value. **d)** The recall, F1 score, AUPRC, false positive rate, NPV, and AUC-ROC under different precisions. The best AUPRC and AUC-ROC are in bold.

### Forecasting high-risk MRSA cases

Identifying MRSA cases in high-risk patients can help target infection control interventions that reduce transmission. Following the approach of Pei et al. [34], we consider a strategy of testing patients based on the ranked carriage probability forecasted by NeurABM and other baselines, and determined the proportion of MRSA cases that can be identified. This is shown in Figure 5, where we rank all patients according to the forecasted infected probability of each method from high to low, and test patients in this order. For example, if we only test 20% of patients based on the ranked risk estimated by NeurABM, we could identify 88% (86 out of 98) of carriage patients. Meanwhile, the best baseline (XGBoost) could only identify 65% (64 out of 98) of them (i.e., 22 less patients than NeurABM). Therefore, NeurABM can always forecast more future nosocomial MRSA infection cases (y-axis) given the same test budget (x-axis), which suggests that our framework is effective and practical in real-world clinical settings.

**Figure 5:**
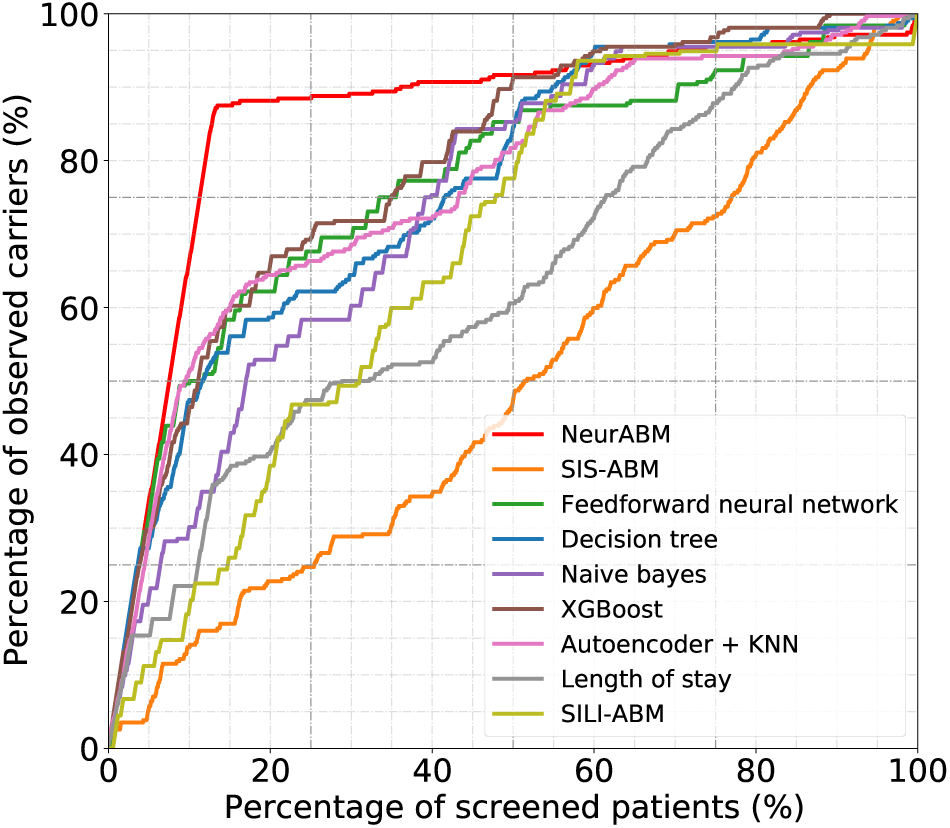
Percentage of identified MRSA cases by screening “high-risk” patients. For each patient in the UVA ICUs, we use each method to estimate their MRSA infection probability and rank them according to this probability from high to low. We then screen different percentages of patients (x-axis) and see how many actual MRSA cases can be forecasted (y-axis). As seen in the figure, NeurABM can always identify more MRSA cases than other baselines.

### Case study: Explanation of neural network

To better demonstrate the interpretability of the NeurABM framework, we further investigated which patient risk factors are considered to have a high risk of being importation cases in the previous results shown in Figure 2 by NeurABM. As shown in Figure 6, each dot represents a patient, and the color of the dot represents the risk factor value (red means higher, and blue means lower). Dots with a higher impact value (to the right of the control line) mean that NeurABM tends to classify this patient as having a higher probability of being an importation case (and vice versa). Here, we can see that patients who have had contact with more MRSA patients in the past 7/14 days prior to ICU admission are considered as having high risk. In addition, we found that patients with a device usage history, and who come from or were discharged to other healthcare facilities last time, are more likely to be considered as importation cases. This also aligns with real-world observations from clinicians [43]. It is important to note that the SHAP values are primarily used to explain the outputs of a neural network, and the results presented in Figure 6 are intended to highlight the interpretability of the NeurABM framework rather than providing direct clinical recommendations or supporting causal conclusions.

**Figure 6:**
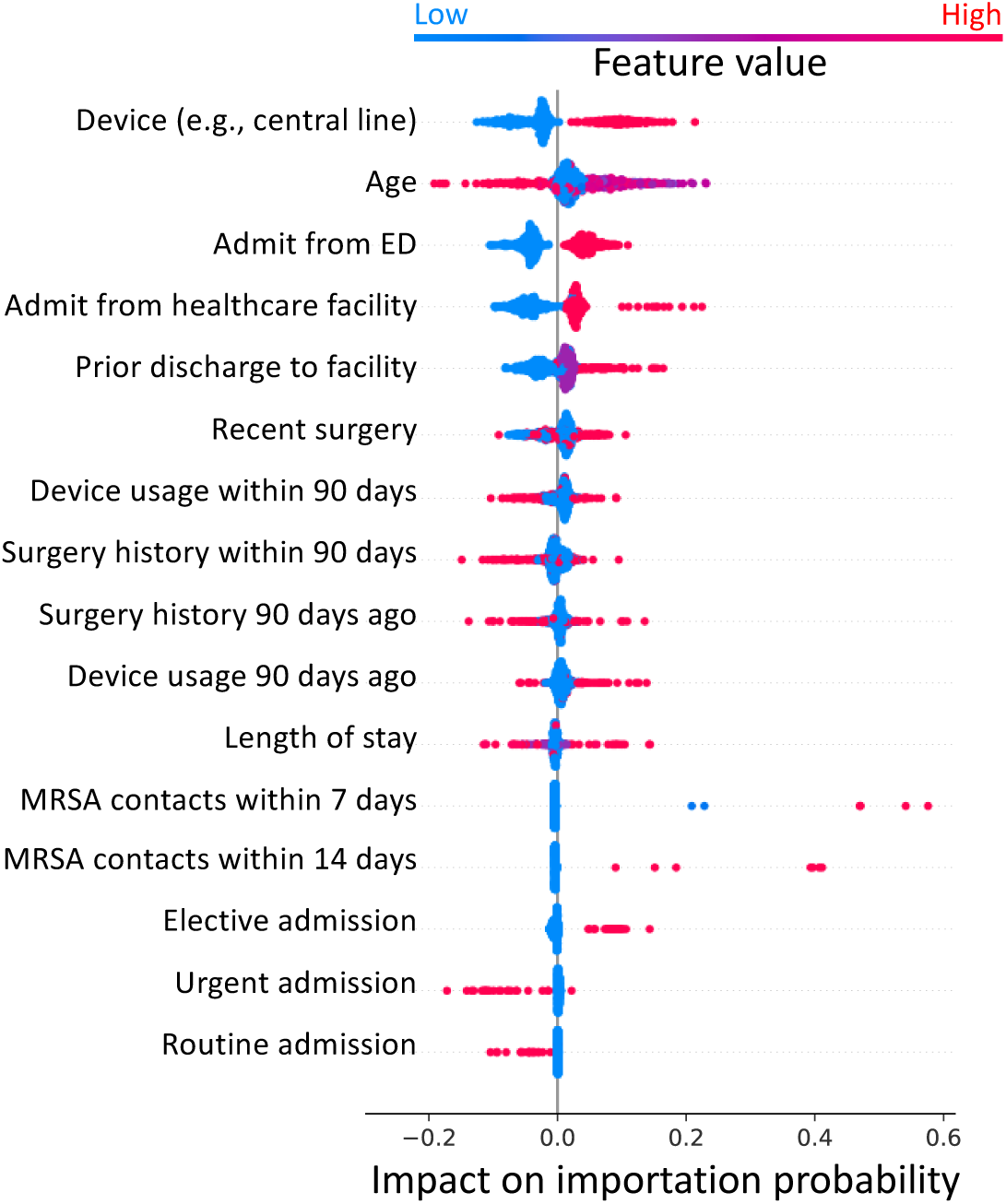
The patient risk factors that are considered as having a high risk of being importation cases by the trained neural network. Color of the dots represent risk factor values, with red indicating higher values. A higher impact means that NeurABM is more likely to consider the patient as an importation case.

## Discussion

Identifying healthcare-associated infections importation and forecasting future nosocomial infection cases have long been a challenging problem. Previous research works relied either solely on machine learning and statistical techniques, which directly use patients’ risk factors collected from EHR data for prediction while overlooking the epidemiological mechanisms involved in HAI spread [37, 22], or solely on mechanistic models that capture the transmission dynamics of HAI within healthcare facilities while ignoring individual patients’ specific risk factors [34]. In contrast, NeurABM integrates both deep learning and ABMs simultaneously in a lock-step to learn importation probabilities. This framework allows the use of diverse data sources from EHRs by deep learning methods and ABMs. Furthermore, the contact network utilized in the SIS-ABM model within NeurABM plays a crucial role and underscores the importance of contact representations in identifying nosocomial infections. Overall, NeurABM is a novel framework for supporting a diverse class of HAI surveillance and control questions.

Our experiments show that NeurABM not only identifies MRSA importation cases and current nosocomial cases but also forecasts future nosocomial infection cases in UVA ICUs, with performance that can be considered clinically useful (e.g., high recall with not-too-low precision as shown in Figure 2a, Figure 3a, and Figure 4a); in contrast, as shown in experiments, prior methods using EHR data haven’t achieved this level of performance. For example, when ranking the patients based on their probability in the carriage state and test the top 20%, we could identify 88% of carriage patients while the best baseline could only identify 65% of them (86 out of 98 vs. 64 out of 98). Moreover, the NPV for our method is always higher than 0.9, indicating that NeurABM can identify and forecast these cases well with fewer missing/undetected patients. Even though importation cases are identified retrospectively, the results remain practical and valuable. Typically, MRSA culture tests require a 3–5 day turnaround time, while the NeurABM framework can provide a rapid estimation during this period, potentially offering actionable insights before culture results are available. Our case study also reveals the risk factors that are highly related to MRSA importations, which confirms clinical observations and allows clinicians to better respond to potential MRSA importation cases in their hospital.

However, our framework is not without limitations. One limitation is that the SIS-ABM model may oversimplify MRSA carriage by defining just two states: *susceptible* and *carriage*. There are different forms of MRSA carriage, including various clinical infection types such as skin abscess or bloodstream infection, which may confer different levels of risk for transmission. Similarly, the SIS-ABM model does not have an explicit “colonization” state. More advanced epidemiological models like multi-strain SIR models could more closely reflect realistic scenarios and further improve performance. Another limitation is that we are currently only using the co-location data to construct the contact networks and assuming that the entities co-located in the same room will have interactions with each other. Recent works have shown that this well-mixed assumption, even for short periods of time or small spatial locations, can be sub-optimal in terms of prediction, while better-designed, norm-based spatial formalization can better capture the interactions between entities in continuous space [26, 25]. Despite the above disadvantages, NeurABM is quite general and can be extended to use more complex ABMs that incorporate these properties separately, depending on the needs in the application and data availability. Besides, MRSA nares testing is widely used in clinical practice to guide decision-making on whether contact precautions and MRSA-active antibiotics should be implemented, and agent-based models (such as the SIS-ABM model used in this paper) are well established for studying MRSA transmission [21, 10]. Therefore, extending agent-based methods to predict MRSA testing outcomes for potential future clinical application is a natural progression of this work, aligning with their established utility in modeling transmission dynamics. Additionally, we are currently evaluating NeurABM only in the UVA ICU due to data availability; adding more patient risk factors (e.g., chronic diseases hisotry or Apache II scores) or conducting more experiments in other settings and healthcare facilities could provide a more comprehensive assessment of NeurABM.

Nevertheless, NeurABM opens up a new direction of research for using both rich patient risk factors from EHR data as well as agent-based models designed with epidemiological knowledge and can be used for many other questions about HAI spread in hospitals. The specific architecture of our method, which runs the deep learning method and ABM step at each time allows us to add predictors at different stages within this framework to address other kinds of questions. For instance, people can also forecast future contact networks instead of naively assuming future contact networks may be the same as previous ones to better forecast future nosocomial infections. The ABM can also be enriched with representations of interventions implemented in the hospital (e.g., how patients are selected and put under contact precautions with identified with MRSA). For patient-specific parameters, we focus on importation probabilities in this work. However, many other parameters such as recovery rate can also be patient-specific and our NeurABM framework can be easily extended to it. The performance of these variations depends on data availability, and is a promising topic for future research.

In terms of real-world applications, the NeurABM framework is highly flexible, as it is not tied to any specific agent-based model. Instead, users can easily replace the SIS-ABM model in NeurABM with their own ABM designs into NeurABM, allowing people to adapt to other healthcare-associated infections, such as *Clostridioides difficile*. Moreover, the datasets required for NeurABM are straightforward to extract from electronic health records [34] (see the Methods section for more details). Our experiments also adhere to a real-world setup: when forecasting future nosocomial infections in week *k*, the model only requires data up to week *k −* 1. This ensures that NeurABM operates without data leakage and is suitable for real-time implementation. The fact that NeurABM is amenable to such adaptations quite easily is an indication of its generality.

## Methods

### Dataset

We extract three different types of patient data based on the electronic health records (EHR) from the University of Virginia hospital: patient demographic information and risk factors (e.g., comorbidities, medical history), lab testing, and contact network data.

- Patient risk factor data: This dataset consists of risk factors for all patients in ICUs. From the EHR dataset, we collected 19 different risk factors for each patient, all of which are available before ICU admission. From July 1, 2019, to December 31, 2019, there were 1,117 patients in UVA ICUs, and 74 of them were MRSA importation cases (all patients received an MRSA test within (*t −* 3*, t* + 3) days of being admitted into one of the ICUs, and patients who tested positive for MRSA within this range are considered as importation cases). A list and description of each risk factor are provided in the Supplementary Information.
- Lab testing data: This dataset consists of infection data for each patient. There were two different types of tests to diagnose MRSA: culture tests and polymerase chain reaction (PCR) tests. However, since a negative culture test cannot disqualify an individual from MRSA infection, we focused on only positive culture tests and both positive and negative PCR tests. For a given patient *p* on a given day *t*, *y_p,t_* = 1 represents that the patient was tested positive on day *t* or if their most recent test in the past was positive. Likewise, *y_p,t_* = 0 if this patient was tested negative on day *t* or if their most recent test was negative.
- Contact network data: This dataset consists of a series of ward-level co-location contact networks ***A****_t_* comprising three different entities: patients, healthcare workers (HCWs), and locations, and each network is for one specific day. From the EHR dataset, we can collect the movement information of patients and HCWs (e.g., the ward that a patient stayed, and when the doctors and nurses visited a specific ward). Note that these movement information also includes start and end times; we can infer whether these patients and HCWs were co-located (i.e., time overlapped) at any specific location. Specifically, if two patients or HCWs *v*_1_, *v*_2_ colocated at location *l* on day *t*, we would create edges between *v*_1_ and *v*_2_, *v*_1_ and *l*, *v*_2_ and *l* on day *t* in ***A****_t_*. However, because of the nature of this data, individuals such as support staff or patient guests are not tracked, and thus are not included in the network. Additionally, HCW-HCW colocations are not tracked in rooms where care is not administered, such as break rooms. From July 1, 2019 to December 31, 2019, there were 1117 patients (and 6445 patient days), 2385 healthcare workers, and 77 locations in the UVA ICU.

### Problem setup

We use the trained NeurABM model to identify importation and nosocomial infection cases. For importation cases, the patient-specific parameter ***θ****_p_* in this work is importation probability for each patient. Specifically, for each week *k*, we used the contact networks, patient risk factors, and lab testing results until week *k* to train the NeurABM, and our task is to identify the importation cases from them until week *k*. Note that NeurABM framework do not access to the ground-truth importation cases data. Instead, we only use the ground-truth importation cases data for evaluation.

For identifying current nosocomial infection cases in hospital, we follow the setup of a previous work [34]: In this setup, since we want to identify current nosocomial infections in week *k*, we should be exactly in week *k* (e.g., at the end of week *k*). Therefore, for each week *k*, we used the contact networks, patient risk factors, and lab testing results until week *k −* 1 to train the NeurABM, and then ran the SIS-ABM model for 7 more days to infer the infection states of all patients for week *k*. Since we are identifying current nosocomial infection cases in week *k*, the contact network in week *k* is known, while the lab test results are not fully available (because of the delays in getting the lab test results). For example, if we were at the end of week 40 (beginning of October), we would train NeurABM on all the data from week 28 (beginning of July) to week 39. As for week 40, we would only use the contact network in that week and use the ABM simulator to identify the nosocomial infection cases in week 40. Then, at the end of week 41, we would train on the data from week 29 to 40 and identify the nosocomial infection cases in week 41. We repeated this procedure until we were at the end of week 52.

For forecasting future nosocomial infection cases, we followed a real-world step-forward scenario that made weekly predictions. In this setup, since we want to forecast future nosocomial infection cases in week *k*, we should be before week *k* (e.g., at the end of week *k −* 1). Therefore, for each week *k*, we used the contact networks, patient risk factors, and lab testing results until week *k −* 1 to train the NeurABM, and then ran the SIS-ABM model for 7 more days to forecast the infection states of all patients for week *k* without knowing any information for week *k*. For example, if we were at the end of week 39, we would train on the data from week 28 to week 39 to forecast for week 40 (i.e., no information from week 40 is used). Then, at the end of week 40, we would train on the data from week 29 to 40 and forecast for week 41. We repeated this procedure until we were at the end of week 52. Intuitively, the key difference between the setup of identifying current nosocomial infection cases and forecasting future nosocomial infection cases is whether the contact network in the targeted week *k* is known or not.

### Transmission model

In this work, we use the SIS-ABM model in NeurABM to capture the MRSA spread dynamics in UVA ICUs [21]. SIS-ABM is a pathogen load-based model that keeps track of pathogen load on all people and locations using a load vector ***l****_t_*. For each patient *i*, they can either be in the *Susceptible* (*S*) or *Carriage* (*C*) state. Specifically, the probability of transitioning from *S* to *C* is proportional to the amount of pathogen on this patient ***l****_t_*(*i*), which can be formulated as a linear dose-response function *β****l****_t_*(*i*) (*β* is the disease infectivity parameter). Once in the carriage state, the patient keeps shedding more pathogen loads at each step, which can later be transferred to its neighbors (including both people and locations). Such a shedding process continues until the patient recovers with a recovery probability *δ*.

For the pathogen load transfer, as described in the previous text, the SIS-ABM model uses daily contact networks ***A****_t_* to capture the exchange of pathogens among patients, HCWs, and locations. Specifically, we construct a transfer matrix ***R****_t_* for each day *t*, where ***R****_ijt_* = *τ_ijt_****A****_ijt_*. Here, ***A****_t_* is the adjacency matrix of contact networks on day *t*, and *τ_ijt_* is the transfer ratio parameter (the ratio of pathogen being transferred (or remaining if *i* = *j*) from patient/HCW/location *j* to patient/HCW/location *i* on day *t*). Specifically, based on the kinds of nodes of *i* and *j*, we have 8 kinds of transfer ratios: *τ_P P_*, *τ_P H_*, *τ_P L_*, *τ_HP_*, *τ_HH_*, *τ_HL_*, *τ_LP_*, *τ_LH_*. It also uses *γ_P_*, *γ_H_*, *γ_L_* to denote the natural pathogen reduction rate on patient, HCW, and location nodes. Using this ***R****_t_* and ***l****_t_*(*i*), the SIS-ABM model updates the pathogen loads every day as a linear operation. We also restrict the column-sums of ***R****_t_* to be less than or equal to 1, which implies that the total amount of pathogen cannot increase after transfer (i.e., *|****R****_t_****l****_t_| ≤ |****l****_t_|*). Note that susceptible patients may still carry a small amount of pathogen loads and spread them to others, and HCWs and locations are always in the *susceptible* state, which means that they can spread the MRSA pathogen loads but are non-infectable. We provide more details in the Supplementary Information.

### NeurABM framework

As shown in Figure 1, the NeurABM framework is composed of two parts: the neural network part (green block, parameterized by *ϕ*) and the agent-based model simulator part (yellow blocks).

For the neural network component, we take the risk factors ***f*** (where *f_p_* is for patient *p*) as input and then estimate both the patient-specific parameters ***θ****_p_* (which is a vector and each element *θ_p_* is for patient *p* and only influenced by the patient itself’s risk factors *f_p_*, in this work it is importation probability for each patient) and ABM parameters ***θ****_M_* (i.e., the general parameters that apply to every patient in the SIS-ABM model, including disease infectivity parameter *β*, recovery probability *δ*, pathogen shedding rate *α*, natural pathogen reduction rate for patients, HCWs, and locations *γ_P_*, *γ_H_*, *γ_L_*, transfer ratios from different kinds of nodes *τ_P P_*, *τ_P H_*, *τ_P L_*, *τ_HP_*, *τ_HH_*, *τ_HL_*, *τ_LP_*, *τ_LH_*) together. The neural network is parameterized by *ϕ* and we use ***θ****_p_,* ***θ****_M_* = *NN* (***f***; *ϕ*) to represent it. We list all ABM parameters in Supplementary Information.

For the ABM simulator, we implement the simulation process of the SIS-ABM model using matrix operations in a differentiable way. ABM simulator takes the adjacency matrices of contact networks ***A****_t_* and the parameters learned by neural networks (***θ****_p_,* ***θ****_M_*) as input, and simulates MRSA spread in *T* days to estimate patient states on each day ***ŷ*** (where *ŷ_p,t_* is for patient *p* on day *t*). Specifically, the simulation process of the SIS-ABM model can be decomposed into three substeps: (1) pathogen load transmission where the load transfers via contact edges, (2) updating the states for each patient based on their pathogen loads and recovery probability, and (3) updating the timestep from day *t* to day *t* + 1. This process can be repeated for arbitrary steps to simulate the MRSA spread over *T* days. We use ***ŷ*** = *ABM* (***A***; ***θ****_p_,* ***θ****_M_*) torepresent it.

We also have another ABM simulator for forecasting. Since we cannot exactly know future contact networks, we assume they will be the same as the contact network on day *T* (i.e., ***A****_T_*). Therefore, it takes the patient states on day *T* as the input and forecast for *T′* days ahead. It can be represented by ***ŷ****′* = *ABM_F_* (***ŷ***; ***A****_T_,* ***θ****_p_,* ***θ****_M_*). However, the assumption on the future contact networks is inaccurate and lead to bias in ***ŷ****′* we forecast. Therefore, we use another adapter neural network *ϕ′* to revise ***ŷ****′*_*F*_ and give our final forecast ***ŷ****_F_*. We formulate it as ***ŷ****_F_* = *Adapter*(***ŷ***′_*F*_, ***f***; *ϕ′*).

With the aforementioned neural network and the ABM simulator, we then integrate them together to train simultaneously. Specifically, one training epoch comprises the following steps.

- Step 1: We feed the risk factor data ***f*** into the neural network as the input to estimate the patient-specific parameters ***θ****_p_*. In this work, ***θ****_p_* is the probability of being importation cases for each patient *p*. It is a vector of size *N*, where *N* is the number of patients in the contact network. Meanwhile, the neural network will also give the general ABM parameters ***θ****_M_* that are applied to all patients (e.g., *β*, *α*, *· · ·*).
- Step 2: We then feed ***θ****_p_*, ***θ****_M_*, and the contact networks ***A****_t_* into the ABM simulator and simulate for *T* steps. The output will be the vector ***ŷ*** of size *N × T*, in which *ŷ_p,t_* represents the probability of being in the state *carriage* for patient *p* on day *t*.
- Step 3: We compare the estimated carriage probability ***ŷ*** with the corresponding ground-truth observations (i.e., known carriage patients based on lab testing) ***y***. We use the weighted binary cross entropy loss (BCE loss) [17] 𝓛(***ŷ***, ***y***) = Ʃ_*p*_ Ʃ_*t*_ *w_pos_y_p,t_* log(*ŷ_p,t_*) + *w_neg_*(1 *− y_p,t_*) log(1 *− ŷ_p,t_*) as the loss function. Here *w_pos_* and *w_neg_* are the weights for positive and negative observations. We set *w_pos_* : *w_neg_* = Ʃ_*p*_ Ʃ_*t*_ 𝟙[*y_p,t_* = 0] : Ʃ_*p*_ Ʃ_*t*_ 𝟙[*y_p,t_* = 1], where 𝟙[*·*] is the indicator function, which is 1 if the condition is true, and 0 otherwise.
- Step 4: Meanwhile, we also feed the *ŷ_p,t_* for day *T* to another ABM simulator to forecast for *T′* days ahead. Note that these *T′* days are for future and we cannot access to the real contact networks when forecasting, we assume that future contact networks will be the same as what we have for day *T*. Let *ABM_F_* be this ABM simulator for forecasting, and ***ŷ′***_*F*_ of size *N × T′* as rough forecast output (in which 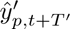 represent the probability of being in the state *carriage* for patient *p* on day *t* + *T′*), we formulate it as ***ŷ′***_*F*_ = *ABM_F_* (***ŷ***; ***A****_T_,* ***θ****_p_,* ***θ****_M_*)
- Step 5: However, the assumption that future contact networks will be the same as on day *T* is inaccurate and leads to bias in 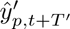. To tackle this, we use another adapter neural network *ϕ′* to revise this rough output to get the revised forecast output ***ŷ****_F_* of size *N ×T′*, in which *ŷ_p,t_*_+_*_T′_* represents the probability of being in the state *carriage* for patient *p* on day *t* + *T′*. We formulate it as ***ŷ***_*F*_ = *Adapter*(***ŷ***′_*F*_, ***f***; *ϕ′*) and compute the BCE loss 𝓛(***ŷ****_F_*, ***y****_F_*) = Ʃ*_p_* Ʃ*_t_ w_pos_y_p,t_*_+_*_T′_* log(*ŷ_p,t_*_+_*_T′_*) + *w_neg_*(1 *−y_p,t_*_+_*_T′_*) log(1 *− ŷ_p,t_*_+_*_T′_*).
- Step 6: With the total loss 𝓛(***ŷ***, ***y***) + 𝓛(***ŷ****_F_*, ***y****_F_*), and the differentiable ABM simulator, we can calculate the gradient of the loss with respect to the neural network parameters *ϕ* and adapter parameters *ϕ′* via backpropagation. This allows us to better tune the neural network and learn more reasonable parameters as the input for the ABM simulator.

The above steps are repeated until the total loss converges. More details are provided in the Supplementary Information.

### Baselines

To compare NeurABM with current modeling or machine learning-based methods, we also compare with other baselines including machine learning-based methods (neural network [33], decision tree [17], naive bayes [6], XGBoost [8], Autoencoder+KNN [36, 15]), mechanistic modeling-based methods (SIS-ABM model [21, 10], SILI-ABM model [34]), and clinical heuristic methods (length of stay [34]).

For machine learning based methods, we train two models: one for identifying importation cases and another for identifying nosocomial infection cases. For importation cases, we train on the ground-truth importation cases from January 2019 to June 2019 and test on July to December (i.e., the same time period for NeurABM). Note that NeurABM identifies importation cases without access to ground-truth data from January 2019 to June 2019. In contrast, the machine learning-based baselines require ground-truth labels as training data. To enable these baselines to function, we provided this additional information, yet they still could not outperform NeurABM. For identifying current and forecasting future nosocomial infection cases in each week *k*, we train on data until week *k −* 1 and test on week *k*. For modeling-based methods, we run their models following their original papers [34, 21] and take the average infected probability of 100 simulations as the probabilities of being importation cases and nosocomial infection cases. For clinical heuristic baselines, the length of stay will consider patients staying longer in the hospital to have higher probabilities.

Since the outputs of NeurABM framework and baseline models are probabilities of being classified as importation or nosocomial infection cases, we applied varying thresholds (ranging from 0 to 1) to convert these probabilities into binary classification outcomes. This allows us to predict whether a patient is an importation or nosocomial infection case when the probability exceeds the threshold, and vice versa. By adjusting the thresholds, we aim to compare different methods more comprehensively. We plotted precision-recall curves (e.g., Figure 2a), ROC curves (e.g., Figure 2c), and the changes in negative predictive value (e.g., Figure 2b) across different thresholds. We also provide a detailed description of the metrics used to evaluate performance in the Supplementary Information.

## Data Availability

The outputs of our model are available on GitHub via link: https://github.com/AdityaLab/NeurABM. The electronic health record (EHR) data used in developing the models is not available since it is highly sensitive, and we do not have permission to release it. However, we provide the code, a demo, and a synthetic dataset on GitHub.

## Inclusion & Ethics statement

This work is a retrospective observational study and follows the STROBE statement checklist [2]. This study was approved by the University of Virginia Institutional Review Board for Health Sciences Research (IRB-HSR-22410), and the written consent was waived due to the retrospective nature of the study.

## Code Availability

The code of our model are also available on GitHub via link: https://github.com/AdityaLab/NeurABM. It also contains a detailed readme file to help run the code.

## Acknowledgements

This work was supported in part by the NSF (Expeditions CCF-1918770, CAREER IIS-2028586, RAPID IIS-2027862, Medium IIS-2403240, Medium IIS-1955883, Medium IIS-2106961, PIPP CCF-2200269), Centers for Disease Control and Prevention Modeling Infectious Diseases In Healthcare program (5U01CK000589), National Institutes of Health (K23AI163368 to G.R.M.) and National Center for Advancing Translational Science (UL1TR003015, KL2TR003016 to G.R.M.), faculty research award from Facebook, Dolby faculty research award, and funds/computing resources from Georgia Tech.

## Author Contributions

J.C., J.H., E.K., G.M., A.V. and B.A.P. conceived the experiments, J.C., and J.H. conducted the experiments, J.C., J.H., E.K., G.M., A.V. and B.A.P. analysed the results. J.C., J.H., E.K., G.M., C.S., A.V. and B.A.P. reviewed the manuscript.

