## Supplementary Information for "Identifying and Forecasting Importation and Asymptomatic Spreaders of Multi-drug Resistant Organisms in Hospital Settings"

### Dataset

We extract three different types of patient data based on the electronic health records (EHR) from the University of Virginia hospital: patient demographic information and risk factors (e.g., comorbidities, medical history), lab testing, and contact network data.

#### Patient risk factor data

This dataset consists of risk factors for patients in intensive care units (ICUs). We extracted 16 different risk factors from the EHR for each patient, which are all available prior to MRSA testing. All patients received an MRSA test within $(t-3, t+3)$ days of being admitted into one of the ICUs, and patients who tested positive for MRSA within this range are considered as importation cases. From July 1, 2019, to December 31, 2019, 1,117 patients spanned the nine different UVA ICU departments. These patients totaled 6,445 patient days, with 74 identified as importation cases. The following provides a definition for each risk factor and the method of their derivation:

- **Device (e.g., central line)**: Binary variable that indicates whether a patient had a catheter, tube, or blood line (defined as "device") administered to them in the UVA hospital system.
- **Age**: Integer value indicating the age of the patient in years.
- **Admit from ED**: Binary variable indicating that the type of admission to the hospital was defined as an emergency.
- **Admit from healthcare facility**: A variable in $-1, 0, 1$ indicating where the patient was admitted from. If the patient was admitted from a skilled nursing facility or a long-term acute care hospital, it is 1; if admitted from somewhere besides these facilities, it is $-1$. It is 0 if the admission location is unknown.
- **Prior discharge to facility**: A variable in $-1, 0, 1$ based on a history of a patient's prior discharges from the UVA hospital system. It is 1 if the patient had previously been discharged to a skilled nursing facility or a long-term acute care hospital and $-1$ if they had been previously discharged but not to one of these facilities. It is 0 if there is no prior discharge data for the patient.
- **Recent surgery**: Binary variable indicating whether the patient has had a surgery performed at any time in the UVA hospital system before the date of the ICU admission.
- **Device usage within 90 days**: Binary variable indicating whether a device has been administered to the patient within the 90 days prior to the ICU admission.
- **Surgery history within 90 days**: Binary variable indicating whether the patient has had a surgery performed in the UVA hospital system within the last 90 days of the ICU admission.
- **Surgery history 90 days ago**: Binary variable indicating if the patient has had surgery in the UVA hospital system more than 90 days prior to the ICU admission.
- **Device usage 90 days ago**: Binary variable indicating whether a device has been administered to the patient more than 90 days prior to the ICU admission.
- **Length of stay**: Integer value defining how long a patient's current stay in the hospital has been before the ICU admission.
- **MRSA contacts within 7 days**: Integer value defining the number of contacts with an individual whose known MRSA status was positive over the 7 days prior to the test administration, as defined by the daily contact networks.
- **MRSA contacts within 14 days**: Integer value defining the number of contacts with an individual whose known MRSA status was positive over the 14 days prior to the test administration, as defined by the daily contact networks.

- **Elective admission**: Binary variable indicating that the type of admission to the hospital was defined as elective.
- **Urgent admission**: Binary variable indicating that the type of admission to the hospital was defined as urgent.
- **Routine admission**: Binary variable indicating that the type of admission to the hospital was defined as routine.

#### Lab testing data

The dataset consists of infection information for each patient. To diagnose MRSA, two types of tests were used in the University of Virginia hospital: culture tests and polymerase chain reaction (PCR) tests. However, a negative culture test does not disqualify an individual from being infected with MRSA, so we concentrated on positive culture tests and both negative and positive PCR tests. For a specific patient $p$ on a specific day $t$, $y_{p,t} = 1$ indicates that the patient tested positive on day $t$ or their most recent previous test was positive. Conversely, $y_{p,t} = 0$ signifies that the patient tested negative on day $t$ or their most recent test was negative.

#### Contact network data

In this work, the agent-based model (ABM) used in NeurABM (SIS-ABM model) also took the contact network data as input. This dataset consists of a series of contact networks comprising three different types of nodes: patients, healthcare workers (HCWs), and locations, and each network is for one day. On a given day $t$, if $i$ and $j$ are colocated at any time on day $t$ in location $l$, an edge will exist between patient/HCW nodes $i$, $j$, and location node $l$. The colocation data is aggregated from the EHR. Specifically, we focus on unweighted and undirected graphs. Therefore, each graph is represented as an $N \times N$ symmetric binary adjacency matrix $\boldsymbol{A}_t$ s.t. $\boldsymbol{A}_t(i,j) = 1$ if nodes $i$ and $j$ are connected with an edge at time $t$, $\boldsymbol{A}_t(i,j) = 0$ otherwise. The node-set is shared across all graphs. Because of the nature of this data, individuals such as support staff or patient guests are not tracked, and thus are not included in the network. Additionally, HCW-HCW colocations are not tracked in rooms where care is not administered, such as break rooms.

### SIS-ABM model

In this work, we use SIS-ABM [11] as the agent-based model in our NeurABM framework to simulate the spread of MRSA. SIS-ABM is "pathogen load-based", where each patient can be either *Susceptible* (S) or *Carriage* (C), and the model keeps track of the pathogen load on all nodes. Such pathogen load-based

Supplementary Table 1: List of SIS-ABM model parameters

| Notation | Description | Range |
|---|---|---|
| $\alpha$ | Pathogen shedding rate | [0,2] [8, 16] |
| $\beta$ | Disease infectivity | [0,0.02] [8, 12] |
| $\delta$ | Recovery probability | [0,0.1] [8, 12] |
| $\gamma_P$ | Natural pathogen reduction rate on patient nodes | [0.9,1] [7] |
| $\gamma_H$ | Natural pathogen reduction rate on HCW nodes | [0.9,1] [7] |
| $\gamma_L$ | Natural pathogen reduction rate on location nodes | [0.9,1] [7] |
| $\tau_{PP}$ | Transfer ratio from patient nodes to patient nodes | [0,0.02] [8, 12] |
| $\tau_{PH}$ | Transfer ratio from patient nodes to HCW nodes | [0,0.02] [8, 12] |
| $\tau_{PL}$ | Transfer ratio from patient nodes to location nodes | [0,0.02] [8, 12] |
| $\tau_{HP}$ | Transfer ratio from HCW nodes to patient nodes | [0,0.02] [8, 12] |
| $\tau_{HH}$ | Transfer ratio from HCW nodes to HCW nodes | [0,0.002] [8, 12] |
| $\tau_{HL}$ | Transfer ratio from HCW nodes to location nodes | [0,0.02] [8, 12] |
| $\tau_{LP}$ | Transfer ratio from location nodes to patient nodes | [0,0.02] [8, 12] |
| $\tau_{LH}$ | Transfer ratio from location nodes to HCW nodes | [0,0.01] [8, 12] |

models are widely used in HAI modeling [17, 5, 15, 4, 19, 13, 2]. We show the pseudocode for the model in Supplementary Algorithm 1. The parameters are listed in the Supplementary Table 1. In SIS-ABM model, each node in the contact networks carries some amount of pathogens that changes over time. The exchange of pathogens among nodes is driven by the edges, which indicates the close contacts between these nodes. It further uses $\tau_{ijt}$ to represent the ratio of the pathogen being transferred from node $j$ to node $i$ at time $t$ (or remaining if $i = j$). Specifically, based on the kinds of nodes of $i$ and $j$, we have 8 kinds of transfer ratios: $\tau_{PP}$, $\tau_{PH}$, $\tau_{PL}$, $\tau_{HP}$, $\tau_{HH}$, $\tau_{HL}$, $\tau_{LP}$, $\tau_{LH}$. It also uses $\gamma_P$, $\gamma_H$, $\gamma_L$ to denote the natural pathogen reduction rate on patient, HCW, and location nodes. Using the transfer ratios $\tau_{ijt}$, reduction rates $\gamma_i$, and adjacency matrix $\boldsymbol{A}_t$, we can construct the transfer matrix $\boldsymbol{R}_t$, and write the pathogen load updates using a linear operation as in Supplementary Algorithm 1 step 5. Here, $\boldsymbol{x}_t$ is the infection state vector at time $t$, $\alpha$ is the pathogen shedding rate for infected patients. Note that the column-sums of $\boldsymbol{R}_t$ are restricted to be less than or equal to 1 (i.e., $|\boldsymbol{R}_t\boldsymbol{l}_t|_1 \leq |\boldsymbol{l}_t|_1$), since the total amount of pathogen cannot increase after transfer. In SIS-ABM model, the probability of a patient in the carriage state increases with the amount of pathogen, which is formulated as a dose-response function $f(\cdot)$ in Supplementary Algorithm 1 step 8. Once infected, the patient sheds $\alpha$ additional pathogen per timestep to his own load, which can later be transferred to neighbors (both people and locations) via edges in contact networks; this shedding continues until the patient recovers. Note that susceptible patients may still be colonized with the pathogen loads and spread them to others. The model also assumes that HCWs and locations that are non-infectable, while they can still act as pathways of pathogen transfer.

**Supplementary Algorithm 1** SIS-ABM model

1: **Inputs:** $\Theta = \left\{\alpha, \beta, \delta, \{\boldsymbol{A}_t\}_{t=1}^{T}, \{\tau_{ijt}\}_{i,j,t}\right\}$
2: Initialize infection-states $\boldsymbol{x}_1$ and loads $\boldsymbol{l}_1$
3: Compute $\boldsymbol{R}_t(i,j) = \begin{cases} \tau_{ijt}\boldsymbol{A}_t(i,j) & \text{if } i \neq j \\ \tau_{ijt}\gamma_i & \text{if } i = j \end{cases}$ for all $t$.
4: **for** $t = 1, \ldots, T$ **do**
5:     Update loads $\boldsymbol{l}_{t+1} = \boldsymbol{R}_t\boldsymbol{l}_t + \alpha\boldsymbol{x}_t$.
6:     **for** each patient $i$ **do**
7:         **if** $i$ is susceptible at time $t$ (i.e., $\boldsymbol{x}_t(i) = 0$) **then**
8:             $i$ gets carriage (i.e. $\boldsymbol{x}_{t+1}(i) = 1$) with probability $f(\boldsymbol{l}_t(i)) = \min\{1, \beta\boldsymbol{l}_t(i)\}$.
9:         **else**
10:             $i$ gets susceptible (i.e. $\boldsymbol{x}_{t+1}(i) = 0$) with probability $\delta$.
11:         **end if**
12:     **end for**
13: **end for**

### NeurABM framework

As shown in Supplementary Figure 1, the NeurABM is composed of two parts: the neural network part (green block, parameterized by $\phi$) and the agent-based model simulator part (yellow blocks).

The neural network part takes the risk factors of patients $\boldsymbol{f}$ (where each element $f_p \in \boldsymbol{f}$ is for patient $p$) as input and then estimates both the patient-specific parameters $\boldsymbol{\theta}_p$ (where $\theta_p \in \boldsymbol{\theta}_p$ is for patient $p$, and in this work it is the probability that one patient is an importation case) and ABM parameters $\boldsymbol{\theta}_M$ (i.e., the general parameters that apply to every patient in the SIS-ABM model, including disease infectivity parameter $\beta$, recovery probability $\delta$, pathogen shedding rate $\alpha$, natural pathogen reduction rate for patients, HCWs, and locations $\gamma_P$, $\gamma_H$, $\gamma_L$, transfer ratios from different kinds of nodes $\tau_{PP}$, $\tau_{PH}$, $\tau_{PL}$, $\tau_{HP}$, $\tau_{HH}$, $\tau_{HL}$, $\tau_{LP}$, $\tau_{LH}$) together. In fact, only learning a good $\boldsymbol{\theta}_M$ is already a non-trivial task in ABM calibration. In this work, we configured the neural network with a depth of 5 layers. Specifically, the neural network part can be represented by

$$\boldsymbol{\theta}_p, \boldsymbol{\theta}_M = NN(\boldsymbol{f}; \phi) \tag{1}$$

The ABM simulator part takes both the parameters from the neural network part (including both patient-specific parameters $\boldsymbol{\theta}_p$ and ABM parameters $\boldsymbol{\theta}_M$) and the adjacency matrices collected from EHR for each day, $\boldsymbol{A}$, as input, and then runs the simulation process as shown in Supplementary Algorithm 1. Specifically,

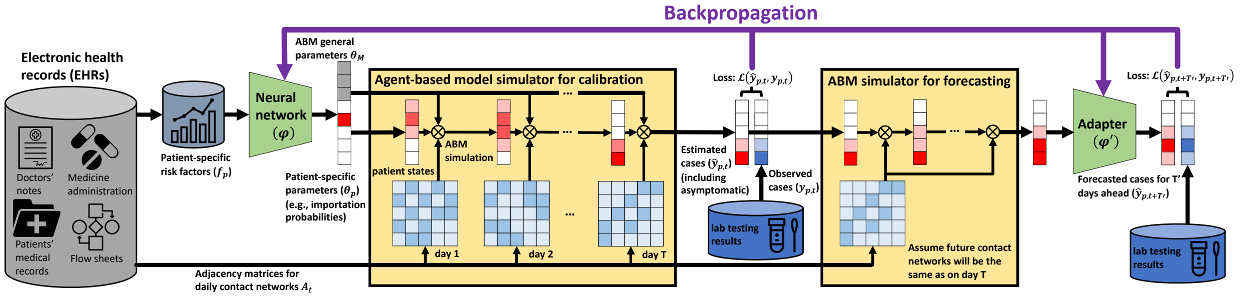


Supplementary Figure 1: **NeurABM framework.** Our NeurABM framework involves 6 steps: (1) The neural network takes the patients' risk factor data collected from EHR as input, and outputs both the agent-based model (ABM) parameters, denoted by $\boldsymbol{\theta}_M$ (which are applicable to every patient), and patient-specific parameters, denoted by $\boldsymbol{\theta}_p$ (importation probabilities in this paper, darker red means higher probability), for each patient $p$. (2) These parameters and the adjacency matrices for contact networks of each day collected from EHR are then fed into the ABM simulator for simulation for $T$ days, and the output will be the probability that each patient $p$ is in the *Carriage* state $\hat{y}_{p,t}$ for each day $t$ (darker red means higher probability). (3) We then compare this $\hat{y}_{p,t}$ with the ground-truth observation of patients in *Carriage* state (via lab testing), denoted by $y_{p,t}$, and compute the loss $\mathcal{L}(\hat{y}_{p,t}, y_{p,t})$. (4) Meanwhile, to forecast future nosocomial infections, we assume that the contact networks will be the same as on day $T$ and further start from $\hat{y}_{p,t}$ on day $T$ and run simulations for $T'$ more days. (5) We also use another adapter network $\phi'$ to correct the potential bias caused by this assumption and get the final forecast $\hat{y}_{p,t+T'}$. We also compute the loss $\mathcal{L}(\hat{y}_{p,t+T'}, y_{p,t+T'})$ with ground-truth observations. (6) With both $\mathcal{L}(\hat{y}_{p,t}, y_{p,t})$ and $\mathcal{L}(\hat{y}_{p,t+T'}, y_{p,t+T'})$, we backpropagate this loss to the neural network $\phi$ and adapter $\phi'$ to tune their parameters.

since the probability that one patient is an importation case is included in $\boldsymbol{\theta}_p$, the patient states at day 0 are known. Other ABM parameters $\boldsymbol{\theta}_M$ are randomly initialized within the range. We calculate the expectations at each time step during the simulation, and the process can be repeated for an arbitrary number of steps to simulate the spread of MRSA over $T$ days. The output is the estimated patient states on each day $\hat{\boldsymbol{y}}$. The ABM part can be represented by

$$\hat{\boldsymbol{y}} = ABM(\boldsymbol{A}; \boldsymbol{\theta}_p, \boldsymbol{\theta}_M) \tag{2}$$

Besides, we also have another ABM simulator for forecasting. Since we cannot exactly know future contact networks, we assume they will be the same as the contact network on day $T$ (i.e., $\boldsymbol{A}_T$). Therefore, it takes the patient states on day $T$ as the input and forecast for $T'$ days ahead. It can be represented by

$$\hat{\boldsymbol{y}}'_F = ABM_F(\hat{\boldsymbol{y}}; \boldsymbol{A}_T, \boldsymbol{\theta}_p, \boldsymbol{\theta}_M) \tag{3}$$

However, the assumption on the future contact networks is inaccurate and lead to bias in $\hat{\boldsymbol{y}}'_F$ we forecast. Therefore, we use another adapter neural network $\phi'$ to revise $\hat{\boldsymbol{y}}'_F$ and give our final forecast $\hat{\boldsymbol{y}}_F$. We formulate it as

$$\hat{\boldsymbol{y}}_F = Adapter(\hat{\boldsymbol{y}}'_F, \boldsymbol{f}; \phi') \tag{4}$$

With both the neural network part and the ABM simulator part, we can then pipeline both parts together to train them simultaneously. Specifically, one training epoch comprises the following steps.

1. **Step 1:** The neural network takes risk factors of each patient as the input to estimate the patient-specific parameters $\boldsymbol{\theta}_p$ and ABM parameters $\boldsymbol{\theta}_M$ as in Supplementary Equation 1. Note that the importation probabilities are also included in the $\boldsymbol{\theta}_p$; thus, we can also get a vector of size $N$ ($N$ is the number of patients in the contact network) that represents the patients' states at $t = 0$.

2. **Step 2:** The ABM simulator then takes both the parameters from the neural network part (including patient-specific parameters $\boldsymbol{\theta}_p$ and ABM parameters $\boldsymbol{\theta}_M$) and the adjacency matrices $\boldsymbol{A}$ collected from EHR for each day as input to run simulations for $T$ steps. The output will be the vector $\hat{\boldsymbol{y}}$ of size

$N \times T$ as in Supplementary Equation 2, where $\hat{y}_{p,t}$ represents the probability of being in state *Carriage* for patient $p$ on day $t$.

3. **Step 3:** We compare the estimated carriage probability $\hat{\boldsymbol{y}}$ with the corresponding ground-truth observations (i.e., observed carriage patients) $\boldsymbol{y}$. More specifically, we use the weighted binary cross-entropy loss (BCE loss) as the loss function.

$$\mathcal{L}(\hat{\boldsymbol{y}}, \boldsymbol{y}) = \sum_{P} \sum_{T} w_{pos} y_{p,t} \log(\hat{y}_{p,t}) + w_{neg}(1 - y_{p,t}) \log(1 - \hat{y}_{p,t}) \tag{5}$$

4. **Step 4:** Meanwhile, the ABM simulator for forecasting takes patients states $\hat{y}_{p,t}$, contact networks for day $T$ ($\boldsymbol{A}_T$), patient-specific parameters $\boldsymbol{\theta}_p$, and ABM parameters $\boldsymbol{\theta}_M$ as input to run simulation for $T'$ more steps for forecasting. The output will be the vector $\hat{\boldsymbol{y}}'_F$ of size $N \times T'$ as in Supplementary Equation 3, where $\hat{y}'_{p,t+T'}$ represents the probability of being in state *Carriage* for patient $p$ on day $t + T'$.

5. **Step 5:** The adapter takes the rough forecast $\hat{y}'_{p,t+T'}$ and the risk factors of each patient $f_p$ as input to revise $\hat{y}'_{p,t+T'}$ and output the final forecast $\hat{\boldsymbol{y}}_F(\hat{y}_{p,t+T'})$, which has the same size and physical meaning as $\hat{\boldsymbol{y}}'_F(\hat{y}'_{p,t+T'})$. To train this adapter, we compare the estimated $\hat{\boldsymbol{y}}_F$ with the corresponding ground-truth observation $\boldsymbol{y}_F$ using the BCE loss as the loss function.

$$\mathcal{L}(\hat{\boldsymbol{y}}_F, \boldsymbol{y}_F) = \sum_{p} \sum_{t} w_{pos} y_{p,t+T'} \log(\hat{y}_{p,t+T'}) + w_{neg}(1 - y_{p,t+T'}) \log(1 - \hat{y}_{p,t+T'}) \tag{6}$$

6. **Step 6:** With the BCE loss $\mathcal{L}(\hat{\boldsymbol{y}}, \boldsymbol{y})$ and $\mathcal{L}(\hat{\boldsymbol{y}}_F, \boldsymbol{y}_F)$, noting that our ABM simulator part is also differentiable as described above, we can then calculate the gradient of the computed loss with respect to each parameter of the neural network via backpropagation. This allows us to better tune the neural network and learn more reasonable parameters as inputs for the ABM simulator.

We repeat the above 6 steps for several epochs until the loss converges. The pseudo code is given in Supplementary Algorithm 2.

**Supplementary Algorithm 2** NeurABM pseudo code

**Inputs:** Risk factors of patients $\boldsymbol{f}$, adjacency matrices of contact networks $\boldsymbol{A}$, lab testing results $\boldsymbol{y}$
**for** each epoch **do**
    Estimate the patient-specific parameters $\boldsymbol{\theta}_p$ and ABM parameters $\boldsymbol{\theta}_M$: $\boldsymbol{\theta}_p, \boldsymbol{\theta}_M = NN(\boldsymbol{f}; \phi)$
    **for** each timestep $t$ **do**
        Estimate the patient states on day $t$: $\hat{\boldsymbol{y}} = ABM(\boldsymbol{A}; \boldsymbol{\theta}_p, \boldsymbol{\theta}_M)$
    **end for**
    **for** each timestep $t$ **do**
        Forecast the patient states on day $t + T'$: $\hat{\boldsymbol{y}}'_F = ABM_F$: $(\hat{\boldsymbol{y}}; \boldsymbol{A}_T, \boldsymbol{\theta}_p, \boldsymbol{\theta}_M)$
    **end for**
    Revise the forecast using adapter: $\hat{\boldsymbol{y}}_F = Adapter(\hat{\boldsymbol{y}}'_F, \boldsymbol{f}; \phi')$
    Compute the loss $\mathcal{L}(\hat{\boldsymbol{y}}, \boldsymbol{y}) + \mathcal{L}(\hat{\boldsymbol{y}}_F, \boldsymbol{y}_F)$
    Update the neural network parameters $\phi$ and adapter parameters $\phi'$
**end for**
**Outputs:** Neural network parameters $\phi$ and adapter parameters $\phi'$

Sometimes, directing the whole NeurABM framework can be extremely hard, since the framework can be really deep. Intuitively, if we use $T$ days for ABM calibration and forecast for $T'$ days ahead, the depth of the framework becomes at least $T + T'$. To address this problem, an iterative training approach following previous work can be helpful [20]: We can iteratively freeze different parts of the frameworks and train the other parts, allowing us to simplify the training NeurABM by breaking it down into two substeps: (1) First, we focus on training the neural network $\phi$ and the ABM simulator $ABM$ while keeping the $\phi'$ and $ABM_F$ frozen, which allows us to first learn reasonable $\boldsymbol{\theta}_M$, $\boldsymbol{\theta}_p$. (2) We then focus on the adapter $\phi'$ and the ABM simulator for forecasting $ABM_F$ while keeping $\phi$ and $ABM$ frozen, which allows us to learn reasonable $\phi'$. These two substeps above can be repeated until convergence.

The formal problem formulation is given as follows:

#### Problem Formulation

Given the risk factors of patients $\boldsymbol{f}$, adjacency matrices of contact networks $\boldsymbol{A}$, lab testing results $\boldsymbol{y}$, the neural network part as in Supplementary Equation 1, the ABM part as in both Supplementary Equation 2 and Supplementary Equation 3, and the adapter network as in Supplementary Equation 4, find the best neural network parameter $\phi$ and adapter neural network parameter $\phi'$ such that

$$\arg\min_{\phi,\phi'} \mathcal{L}(\hat{\boldsymbol{y}}, \boldsymbol{y}) + \mathcal{L}(\hat{\boldsymbol{y}}_F, \boldsymbol{y}_F) \tag{7}$$

With the trained NeurABM model, we can identify importation and nosocomial infection cases. For importation cases, note that the neural network part can estimate both patient-specific parameters $\boldsymbol{\theta}_p$ and ABM parameters $\boldsymbol{\theta}_M$. We can just use this $\boldsymbol{\theta}_p$ as the estimated probability of being an importation case for each patient.

#### Baselines

To compare our NeurABM with current modeling or machine learning-based methods, we also compare with other baselines including Feedforward neural network, decision trees, naive bayes, SIS-ABM, SILI-ABM, clinical heuristic methods, and so on. For machine learning-based methods, we train two models: one for identifying importation cases, and another for identifying nosocomial infection cases.

- SIS-ABM: As an ablation study, we use only the SIS-ABM model to identify importation and nosocomial infection cases to see how it performs.
- SILI-ABM: Sequential, individual-level inference (SILI) model [15] is an agent-based model that uses Bayesian approaches to infer the colonization probability of confirmed carriers for each patient.
- Feedforward neural network: Feedforward neural networks have shown good performance in different clinical estimation and prediction tasks in recent years. In this work, we also implemented a multi-layer perceptron (MLP) feedforward neural network using Scikit-learn [14] in Python.
- Decision tree: Decision tree is also a widely-used machine learning algorithm in classification and prediction [3]. It represents choices in a tree-like graph, illustrating different outcomes as branches. In this work, we also use a decision tree as a baseline using Scikit-learn [14] in Python.
- Naive bayes: Naive bayes is also a powerful and efficient algorithm in machine learning [10]. It operates on the principle of Bayes theorem, where "naive" assumes that features are independent. Similarly, we use Scikit-learn [14] and implemented a naive bayes classifier.
- XGBoost: XGBoost (Extreme Gradient Boosting) is another powerful, scalable machine learning algorithm based on decision trees. It is designed for speed, accuracy, and efficiency, excelling in structured/tabular data predictive tasks [6]. We use the XGBoost Python package [1] and implemented an XGBoost classifier.
- Autoencoder+KNN: We also implemented another machine learning baseline that integrates both a fully connected autoencoder with the K Nearest Neighbors (KNN) algorithm following previous works [18, 9]. Specifically, we first use an autoencoder to learn the latent representation for each patient based on their risk factors, and then use Scikit-learn [14] to implement a KNN classifier.
- Length of stay: As suggested by a previous study [15], patients with longer hospital stay days are suspected of having a higher risk of infection. Therefore, we use the EHR to calculate the days a patient stays in the hospital, and use it as the clinical heuristic baseline for comparison.

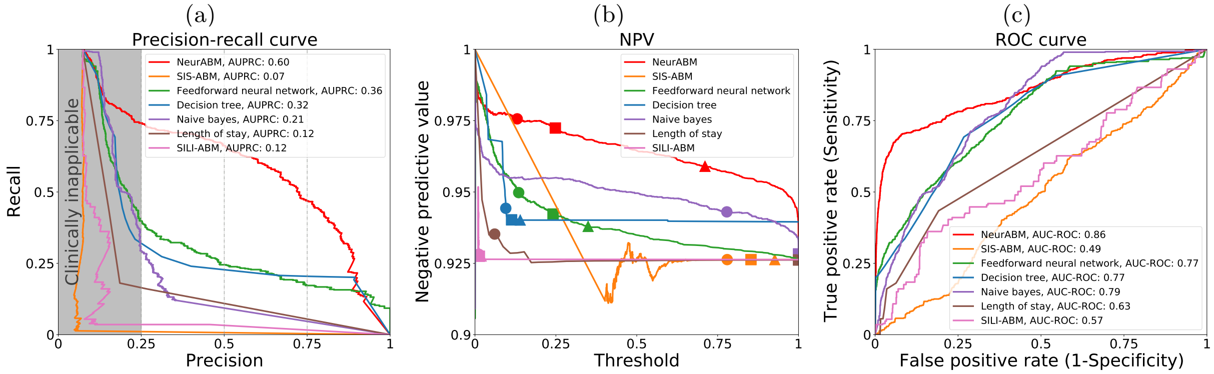


(d) Identifying MRSA importation cases

| Method | Precision | Recall (Sensitivity, True positive rate) | F1 score | AUPRC | False positive rate (1-Specificity) | Negative predictive value (NPV) | AUC-ROC |
|---|---|---|---|---|---|---|---|
| NeurABM | 0.25<br>0.50<br>0.75 | 0.74<br>0.66<br>0.47 | 0.37<br>0.57<br>0.58 | **0.60** | 0.18<br>0.05<br>0.01 | 0.98<br>0.97<br>0.96 | **0.86** |
| SIS-ABM | 0.25<br>0.25<br>0.75 | 0.01<br>0.01<br>0.01 | 0.02<br>0.01<br>0.01 | 0.07 | 0.01<br>0.01<br>0.01 | 0.93<br>0.93<br>0.93 | 0.49 |
| Feedforward neural network | 0.25<br>0.50<br>0.75 | 0.40<br>0.24<br>0.17 | 0.31<br>0.33<br>0.28 | 0.36 | 0.10<br>0.02<br>0.01 | 0.95<br>0.94<br>0.94 | 0.77 |
| Decision tree | 0.25<br>0.50<br>0.75 | 0.32<br>0.23<br>0.20 | 0.28<br>0.31<br>0.32 | 0.32 | 0.08<br>0.02<br>0.01 | 0.94<br>0.94<br>0.94 | 0.77 |
| Naive bayes | 0.25<br>0.50<br>0.75 | 0.29<br>0.09<br>0.05 | 0.27<br>0.16<br>0.09 | 0.21 | 0.07<br>0.02<br>0.01 | 0.94<br>0.93<br>0.93 | 0.79 |
| XGBoost | 0.25<br>0.50<br>0.75 | 0.39<br>0.28<br>0.26 | 0.30<br>0.36<br>0.38 | 0.40 | 0.09<br>0.02<br>0.01 | 0.95<br>0.94<br>0.94 | 0.83 |
| Autoencoder+KNN | 0.25<br>0.50<br>0.75 | 0.01<br>0.01<br>0.01 | 0.01<br>0.01<br>0.01 | 0.10 | 0.01<br>0.01<br>0.01 | 0.93<br>0.93<br>0.93 | 0.62 |
| Length of stay | 0.25<br>0.50<br>0.75 | 0.16<br>0.11<br>0.05 | 0.20<br>0.18<br>0.10 | 0.12 | 0.04<br>0.03<br>0.02 | 0.94<br>0.93<br>0.93 | 0.63 |
| SILI-ABM | 0.25<br>0.50<br>0.75 | 0.04<br>0.03<br>0.02 | 0.06<br>0.06<br>0.03 | 0.12 | 0.02<br>0.01<br>0.01 | 0.93<br>0.93<br>0.93 | 0.57 |

Supplementary Figure 2: **The performance in identifying importation cases. a)** The precision-recall curves (PRC). The x-axis represents precision, and the y-axis represents recall. The red and other color curves represent NeurABM and other baselines. A larger area under the precision-recall curve (AUPRC) indicates better performance. AUPRC values are listed in the legends, and NeurABM has the highest AUPRC value. **b)** The negative predictive value (NPV) with different thresholds. The x-axis is the threshold for classification, and the y-axis is the NPV value. Circles, squares, and triangles correspond to the thresholds and NPV values where precision is 0.25, 0.5, and 0.75, respectively. A higher NPV value indicates fewer missing importation cases that are not identified and therefore better performance, and NeurABM has the highest NPV values. **c)** The receiver operating characteristic (ROC) curves in identifying MRSA importation cases. The x-axis is the false positive rate, and the y-axis is the true positive rate. A larger area under the ROC (AUC-ROC) indicates better performance. AUC-ROC values are listed in the legends, and NeurABM has the highest AUC-ROC value. **d)** The recall, F1 score, AUPRC, false positive rate, NPV, and AUC-ROC under different precisions (0.25, 0.5, 0.75). The best AUPRC and AUC-ROC are in bold.

### Metrics

To evaluate the performance of our NeurABM and other baselines, we use the following metrics in the article:

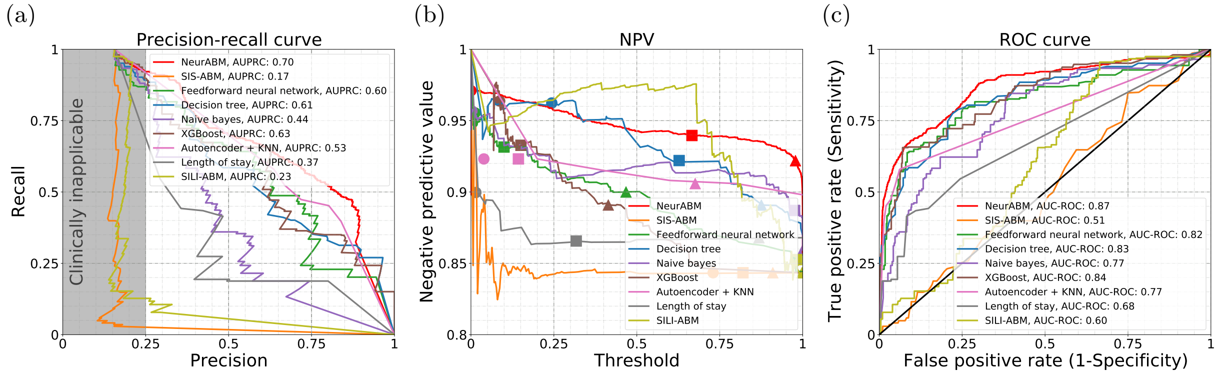


(d) Forecasting future nosocomial infection cases

| Method | Precision | Recall (Sensitivity, True positive rate) | F1 score | AUPRC | False positive rate (1-Specificity) | Negative predictive value (NPV) | AUC-ROC |
|---|---|---|---|---|---|---|---|
| NeurABM | 0.25<br>0.50<br>0.75 | 0.92<br>0.70<br>0.56 | 0.39<br>0.58<br>0.64 | **0.70** | 0.52<br>0.13<br>0.03 | 0.97<br>0.94<br>0.92 | **0.87** |
| SIS-ABM | 0.25<br>0.50<br>0.75 | 0.03<br>0.02<br>0.01 | 0.05<br>0.03<br>0.02 | 0.17 | 0.01<br>0.01<br>0.01 | 0.84<br>0.84<br>0.84 | 0.51 |
| Feedforward neural network | 0.25<br>0.50<br>0.75 | 0.87<br>0.65<br>0.24 | 0.39<br>0.57<br>0.37 | 0.60 | 0.55<br>0.12<br>0.02 | 0.96<br>0.93<br>0.90 | 0.82 |
| Decision tree | 0.25<br>0.50<br>0.75 | 0.89<br>0.59<br>0.36 | 0.39<br>0.54<br>0.48 | 0.61 | 0.51<br>0.12<br>0.02 | 0.96<br>0.92<br>0.89 | 0.83 |
| Naive bayes | 0.25<br>0.50<br>0.75 | 0.87<br>0.36<br>0.10 | 0.39<br>0.42<br>0.18 | 0.44 | 0.50<br>0.08<br>0.01 | 0.96<br>0.89<br>0.86 | 0.77 |
| XGBoost | 0.25<br>0.50<br>0.75 | 0.90<br>0.66<br>0.36 | 0.39<br>0.57<br>0.48 | 0.63 | 0.51<br>0.15<br>0.02 | 0.96<br>0.93<br>0.89 | 0.84 |
| Autoencoder+KNN | 0.25<br>0.50<br>0.75 | 0.92<br>0.70<br>0.47 | 0.39<br>0.58<br>0.57 | 0.53 | 0.82<br>0.33<br>0.03 | 0.92<br>0.92<br>0.91 | 0.77 |
| Length of stay | 0.25<br>0.50<br>0.75 | 0.69<br>0.19<br>0.19 | 0.37<br>0.28<br>0.30 | 0.37 | 0.48<br>0.05<br>0.04 | 0.90<br>0.87<br>0.87 | 0.68 |
| SILI-ABM | 0.25<br>0.50<br>0.75 | 0.13<br>0.06<br>0.03 | 0.17<br>0.10<br>0.05 | 0.23 | 0.11<br>0.01<br>0.01 | 0.85<br>0.85<br>0.84 | 0.60 |

Supplementary Figure 3: **The performance in forecasting future nosocomial infection cases. a)** The precision-recall curves. The red and other color curves represent NeurABM and other baselines. Higher AUPRC is better, and NeurABM has the highest AUPRC value. **b)** The negative predictive value with different thresholds. Circles, squares, and triangles correspond to the thresholds and NPV values where precision is 0.25, 0.5, and 0.75, respectively. Higher NPV value is better, and NeurABM has the highest NPV values. **c)** The receiver operating characteristic curves in forecasting future MRSA nosocomial infection cases. Higher AUC-ROC is better, and NeurABM has the highest AUC-ROC value. **d)** The recall, F1 score, AUPRC, false positive rate, NPV, and AUC-ROC under different precisions. The best AUPRC and AUC-ROC are in bold.

- Precision [10]: Precision measures the proportion of true positive cases (predicted positive and ground-truth positive) among all predicted positive cases, which indicates how many predicted positive cases are ground-truth positive. Higher is better.

- Recall (sensitivity, true positive rate) [10]: Recall measures the proportion of true positive cases among ground-truth positive cases, which indicates how many ground-truth positive cases are predicted as

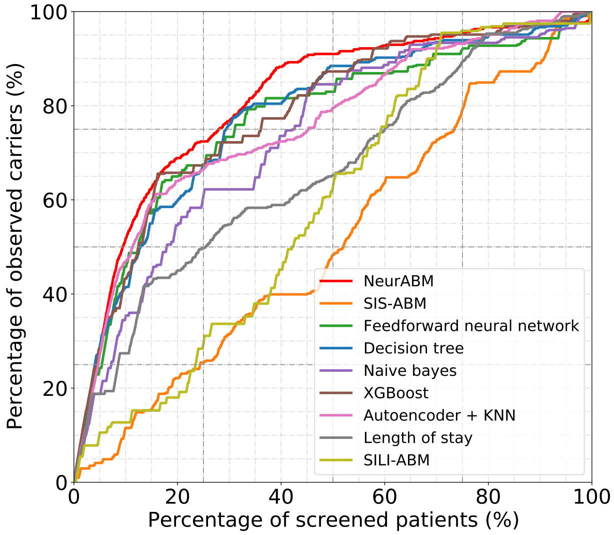


Supplementary Figure 4: **Percentage of identified MRSA cases by screening "high-risk" patients.** For each patient in the UVA ICUs, we use each method to estimate their MRSA infection probability and rank them according to this probability from high to low. We then screen different percentages of patients (x-axis) and see how many actual MRSA cases can be forecasted (y-axis). As seen in the figure, NeurABM can always identify more MRSA cases than other baselines.

positive (i.e., the ability to find all ground-truth positive cases). Higher is better.

- F1 score [10]: F1 score combines both precision and recall into a single measure, which is the harmonic mean of precision and recall (i.e., $\frac{2}{\frac{1}{\text{precision}}+\frac{1}{\text{recall}}}$). Higher is better.
- Precision-recall curve [14]: The precision-recall curve shows the trade-off between precision and recall for different thresholds.
- AUPRC [14]: The area under the precision-recall curve. A high AUPRC value represents both high recall and high precision, and therefore indicates better performance.
- False positive rate (1-specificity) [3]: False positive rate measures the proportion of false positive cases (predicted positive but ground-truth negative) among all ground-truth negative cases, which indicates how many ground-truth negative cases are wrongly predicted as positive. Lower is better.
- Negative predictive value (NPV) [3]: Negative predictive value measures the proportion of true negative cases (predicted negative and ground-truth negative) among all predicted negative cases, which indicates how many predicted negative cases are ground-truth negative. Higher is better.
- Receiver operating characteristic curve [14]: The receiver operating characteristic curve shows the trade-off between true positive rate (recall or sensitivity) and false positive rate (1-specificity) for different thresholds.
- AUC-ROC [10, 14]: The area under the receiver operating characteristic curve. A high AUC-ROC value means a higher probability of predicting ground-truth positive cases as positive compared to ground-truth negative cases, and therefore indicates better performance.

### Additional experiment results

In addition to the baselines listed in the main article, we also compared with SILI model [15] in identifying importation cases. As shown in Supplementary Figure 2, our NeurABM (red) achieves better performance than the SILI model (pink).

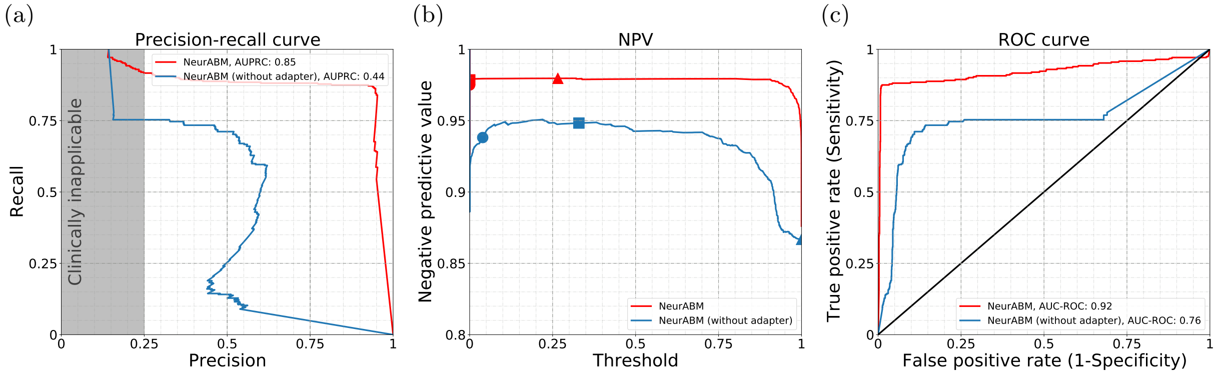


(d) Ablation study: Forecasting future nosocomial infection cases

| Method | Precision | Recall (Sensitivity, True positive rate) | F1 score | AUPRC | False positive rate (1-Specificity) | Negative predictive value (NPV) | AUC-ROC |
|---|---|---|---|---|---|---|---|
| NeurABM | 0.25 | 0.92 | 0.39 | **0.85** | 0.46 | 0.97 | **0.92** |
| | 0.50 | 0.89 | 0.64 | | 0.20 | 0.98 | |
| | 0.75 | 0.88 | 0.81 | | 0.10 | 0.98 | |
| NeurABM (without adapter) | 0.25 | 0.75 | 0.38 | 0.44 | 0.68 | 0.94 | 0.76 |
| | 0.50 | 0.71 | 0.59 | | 0.13 | 0.95 | |
| | 0.75 | 0.05 | 0.09 | | 0.01 | 0.87 | |

Supplementary Figure 5: **Ablation study: The performance in forecasting future nosocomial infection cases of NeurABM and NeurABM (without adapter) a)** The precision-recall curves. The red and blue curves represent NeurABM and NeurABM (without adapter). Higher AUPRC is better,. **b)** The negative predictive value with different thresholds. Circles, squares, and triangles correspond to the thresholds and NPV values where precision is 0.25, 0.5, and 0.75, respectively. Higher NPV value is better. **c)** The receiver operating characteristic curves in forecasting future MRSA nosocomial infection cases. Higher AUC-ROC is better. **d)** The recall, F1 score, AUPRC, false positive rate, NPV, and AUC-ROC under different precisions. The best AUPRC and AUC-ROC are in bold.

We also ran the experiments to forecast for 14 days ahead, and the results show that NeurABM still performs better than other baselines. As shown in Supplementary Figure 3a, the x-axis and y-axis represent precision and recall, respectively. The red curve represents our NeurABM framework. As shown in the figure, the area under the precision-recall curve for our framework is the largest (0.70) compared to other baselines. The dashed grey lines correspond to the precision of 0.25, 0.5, and 0.75. Again, our NeurABM always achieves the highest recall with precision equal to 0.25, 0.5, and 0.75, indicating that our framework is effective. In Supplementary Figure 3b, we show how the negative predictive value (NPV) changes with threshold changes. We can see that the NPV rate is always higher than 0.9 and other baselines, indicating that our NeurABM can forecast future nosocomial infection cases well with fewer missing/undetected patients. We also show the ROC curve in Supplementary Figure 3c. Here, the area under the ROC curve for our framework is the largest (0.87) compared to other baselines. In the table in Supplementary Figure 3d, NeurABM always achieves the highest recall and F1 score with a given precision, demonstrating the effectiveness of our framework in forecasting future nosocomial MRSA infection cases.

To better examine the performance of NeurABM and other baselines for infection control within the hospital, following the setup of previous study [15], we consider the strategy of testing patients based on the ranked carriage probability forecasted by NeurABM and other baselines, and determine what percentage of MRSA cases can be identified. As shown in Supplementary Figure 4, we rank all patients according to the forecasted infected probability of each method from high to low, and test patients in this order. Here we can see that our NeurABM can always forecast more future nosocomial MRSA infection cases (y-axis) given the same test budget (x-axis), which suggests that our framework is effective and practical in real-world clinical settings.

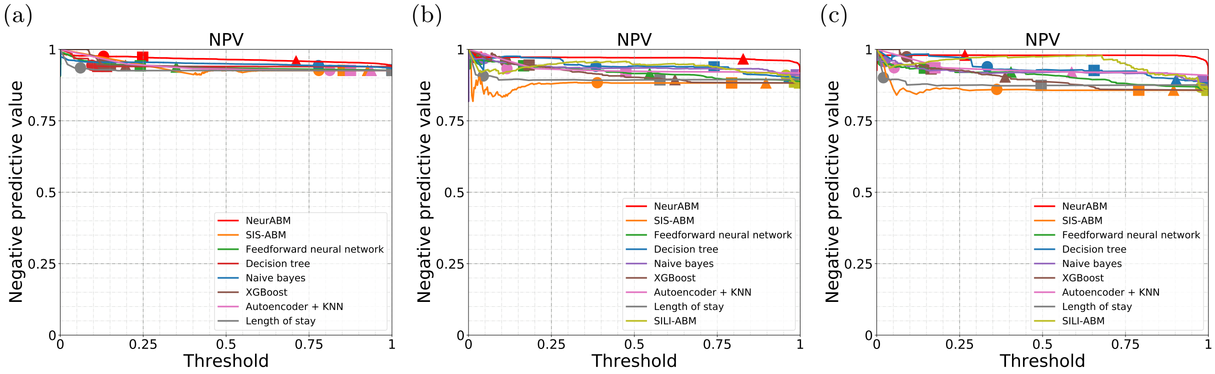


Supplementary Figure 6: **The negative predictive value (NPV) for different tasks.** The x-axis is the threshold for classification, and the y-axis is the NPV value ranging from 0 to 1. Circles, squares, and triangles correspond to the thresholds and NPV values where precision is 0.25, 0.5, and 0.75, respectively. A higher NPV value indicates better performance, and NeurABM has the highest NPV values for all tasks. **a)** for identifying importation cases (i.e., Figure 2b in main article) **b)** for identiying current nosocomial infection cases (i.e., Figure 3b in main article), **c)** for forecasting future nosocomial infection cases (i.e., Figure 4b in main article).

### Ablation study

We also added an ablation study to show the effectiveness of the adapter network $\phi'$. Specifically, we show how the rough forecast $\hat{\boldsymbol{y}}'_F$ and the revised forecast $\hat{\boldsymbol{y}}_F$ before and after the adapter $\phi'$ perform in forecasting future nosocomial infection cases. As shown in Supplementary Figure 5, NeurABM achieves higher AUPRC and AUC-ROC than NeurABM without adapter, showing that the adapter $\phi'$ does help NeurABM forecast better future nosocomial infections.

### NPV rate

To better showcase the NPV rate for identifying importation cases, current nosocomial infection cases, and forecasting future nosocomial infection cases, we also show Figure 2b, 3b, and 4b in the main article by changing the y-axis range from 0 to 1 as in Supplementary Figure 6.
